# Prospective validation of a seizure diary forecasting falls short

**DOI:** 10.1101/2024.01.11.24301175

**Authors:** Daniel M. Goldenholz, Celena Eccleston, Robert Moss, M. Brandon Westover

**Author notes:** Corresponding author: Daniel Goldenholz, 330 Brookline Ave Baker 5, Boston MA 02215.

## Abstract

**OBJECTIVE:** Recently, a deep learning AI model forecasted seizure risk using retrospective seizure diaries with higher accuracy than random forecasts. The present study sought to prospectively evaluate the same algorithm.

**METHODS:** We recruited a prospective cohort of 46 people with epilepsy; 25 completed sufficient data entry for analysis (median 5 months). We used the same AI method as in our prior study. Group-level and individual-level Brier Skill Scores (BSS) compared random forecasts and simple moving average forecasts to the AI.

**RESULTS:** The AI had an AUC of 0.82. At the group level, the AI outperformed random forecasting (BSS=0.53). At the individual level, AI outperformed random in 28% of cases. At the group and individual level, the moving average outperformed the AI. If pre-enrollment (non-verified) diaries (with presumed under-reporting) were included, the AI significantly outperformed both comparators. Surveys showed most did not mind poor quality LOW-RISK or HIGH-RISK forecasts, yet 91% wanted access to these forecasts.

**SIGNIFICANCE:** The previously developed AI forecasting tool did not outperform a very simple moving average forecasting this prospective cohort, suggesting that the AI model should be replaced.

**Key points:** A previously developed e-diary based AI seizure forecasting tool was prospectively tested. Although by some metrics the tool was successful, the overall AI performance was unacceptably low.

It was much easier to outperform a random forecast; it was much harder to outperform a moving average forecast.

Using unverified diaries can skew forecasting metrics in favor of underperforming tools.

## Introduction

Not knowing when the next seizure will happen reduces quality of life for people living with epilepsy. Roughly a decade ago, it was discovered that it is possible to provide seizure forecasts using invasive technology^1^. Since then, novel approaches involving highly invasive^2–5^ and less invasive tools^6,7^ have been proposed. Using a retrospective study of 5,419 unverified self-reported electronic diaries from Seizure Tracker, our group reported that 24-hour forecasts from seizure diaries alone were possible using deep learning^8^. The present study aimed to validate these findings prospectively.

## Methods

### Patients

The protocol was deemed Exempt by the BIDMC Institutional Review Board. Participants were recruited by Seizure Tracker^9^ via email. Participants with 1) epilepsy, 2) age 18 or older, 3) an active Seizure Tracker e-diary account, 4) at least 3 seizures recorded in their account, and 5) at least 3 months of previous e-diary data were eligible. Verified participants linked their e-diary and a RedCap^10,11^ survey account to the study. They completed an initial survey and then weekly surveys (verifying diary completion) for 5 months. They also maintained seizure e-diaries. For safety, only retrospective forecasts were provided monthly.

### The AI forecaster

Using our pre-trained deep learning algorithm^8^ (hereafter: AI), seizure forecasts were calculated for every day possible. The AI uses a recurrent neural network connected to a multilayer perceptron trained on 3806 users (Appendix A). All model parameters and hyperparameters remained unchanged from the original model.

The AI computes a probability of any seizures occurring within a 24-hour period. The AI uses the 84-day trailing history of daily seizure counts leading up to that forecasted day as input. The tool was applied sequentially with a sliding window that moves forward one day at a time. Each patient could have up to 57 daily forecasts (8 weeks and one day), representing the prospective observation period. In some patients, this number was lower due to incomplete diary information (Appendix B). The 3-month pre-enrollment diaries were retained for additional analysis.

### The random forecaster

The daily AI forecast was compared with a permuted forecaster as a benchmark (hereafter “random”). The random forecaster is generated by permuting forecasts from the AI at the subject level. This can be thought of as shuffling a deck of cards, where each card is the AI forecast for a given day, and there is a different deck for each patient. A useful forecast should (at minimum) outperform a permuted forecaster^12^. Where appropriate, the average outcome metric from 1000 such permutations was used, such as for computing the Brier Score.

### The moving average forecaster

The daily AI forecast was also compared with a moving average forecaster which accounted for the typical seizure rate from each patient. Moving average forecasts were computed by taking the total number of seizure days in each trailing 84-day history and dividing by 84 to obtain a simple estimate of daily risk of any seizures for the coming 24-hour forecast (Appendix A). Of note, unlike a similar comparator used our prior study (there called the “rate matched random” forecaster), this moving average forecaster uses total seizure days, not total seizure counts^8^. This change was made to provide a more stringent comparator for the AI. Also of note, all summary results were computed using only the verified post-enrollment period due to concerns about possible under-reporting during the pre-enrollment period (see Discussion).

### Outcome metrics

Performance of each model was measured using area under the receiver operating characteristic curve (AUC), and the Brier Score. AUC values range between 0 and 1, with 0.5 representing a tool indistinguishable from coin flipping, and 1 representing a perfect discriminator. Brier Scores range between 0 and 1, with values closer to 0 representing higher accuracy. Our primary outcome (Appendix B) was comparing AI to the random forecasts using Brier Skill Scores (BSS). Brier Skill Score of 1 represents the AI algorithm is perfect, 0 indicates the AI is not better than the reference forecast, and −1 indicates the reference forecast is perfect).

BSS was computed both at the group-level and at the individual participant level. When using as reference test the random forecaster to calculate BSS, “group-level” means that random forecasts were generated by randomly shuffling the AI predictions across all patients, and randomly reassigning them. Note that this means that forecasts from one patient may be randomly reassigned to other patients. By contrast, calculating BSS at the “individual level” relative to random forecasting means that random forecasts are all from the same patients, albeit in a randomly shuffled order. This means that the group and individual level BSS scores are not directly comparable, and the median of the individual-level BSS scores need not match the group-level BSS score. Additional BSS values were computed using the moving average as an alternative reference.

Calibration curves were generated for the AI, random, and moving average forecasters using equally spaced bins. Confidence intervals for AUC and BSS values were obtained by 1000 bootstrapped samples, selecting patients with replacement.

Code is available here: https://github.com/GoldenholzLab/deepManCode.

## Results

Of 46 recruited participants, 1 was ineligible, 3 were seizure-free, and 11 provided insufficient diary data. Within the remaining 31, there were 3 dropouts, and 8 who missed some of the weekly diary completeness responses. Only 25 patients had sufficient contiguous data to perform forecasts based on 3 months of prospectively collected history. Forecastable diary days (Appendix C) ranged 15-57 (median 57) days. Total seizures per patient ranged from 1-56, (median 13). Participant characteristics are summarized in Table 1.

**TABLE 1:** Baseline characteristics of participants in the prospective study. Note, 31 patients had sufficient information to proceed to analysis, however 6 did not have sufficient data for analysis involving forecasts made only from 3 months of prospectively collected history.

|  | Number | % |
| --- | --- | --- |
| Number of patients | 31 |  |
| Females (%) | 14 | 45% |
| Physician confirmed epilepsy | 31 | 100% |
| EEG confirmed epilepsy |  |  |
| Yes | 27 | 87% |
| Unsure | 4 | 13% |
| Handedness (right / left / mixed) |  |  |
| Right | 23 | 74% |
| Left | 6 | 19% |
| Mixed | 2 | 6% |
| Epilepsy type |  |  |
| Generalized | 8 | 26% |
| Focal | 11 | 35% |
| Focal + Generalized | 8 | 26% |
| Unknown | 4 | 13% |
| Epilepsy location |  |  |
| Frontal | 1 | 3% |
| Temporal | 6 | 19% |
| Parietal | 0 | 0% |
| Occipital | 1 | 3% |
| Multifocal | 2 | 6% |
| Unknown | 21 | 68% |
| Epilepsy cause |  |  |
| Structural | 9 | 29% |
| Genetic | 6 | 19% |
| Infectious | 1 | 3% |
| Metabolic | 0 | 0% |
| Immune | 0 | 0% |
| Unknown | 15 | 48% |
| Prior epilepsy surgery (%) | 16 | 52% |

### Group level results

The following represent group level metrics (Figure 1). Confidence intervals were obtained via 1000 bootstrapped samples with replacement at the patient level. The AUC for AI was: 0.82 [95% CI 0.72-0.90], and for the permuted AI (i.e. random forecast) was 0.50 [95% CI 0.46-0.54]. The Brier Score for AI was 0.14. The AI performed significantly better than the random forecaster at the group level, with a Brier Skill Score (AI vs. random) of 0.53 [95% CI 0.27-0.70]. However, the AUC of the moving average forecaster was also 0.82 [95% CI 0.72-0.89], which was not significantly different from the AI (Mann Whitney U, p=0.13); and the Brier Skill Score of the AI relative to the moving average forecaster was −0.01 [95% CI −0–04 − 0.02], suggesting minimal difference in performance.

**Figure 1:**
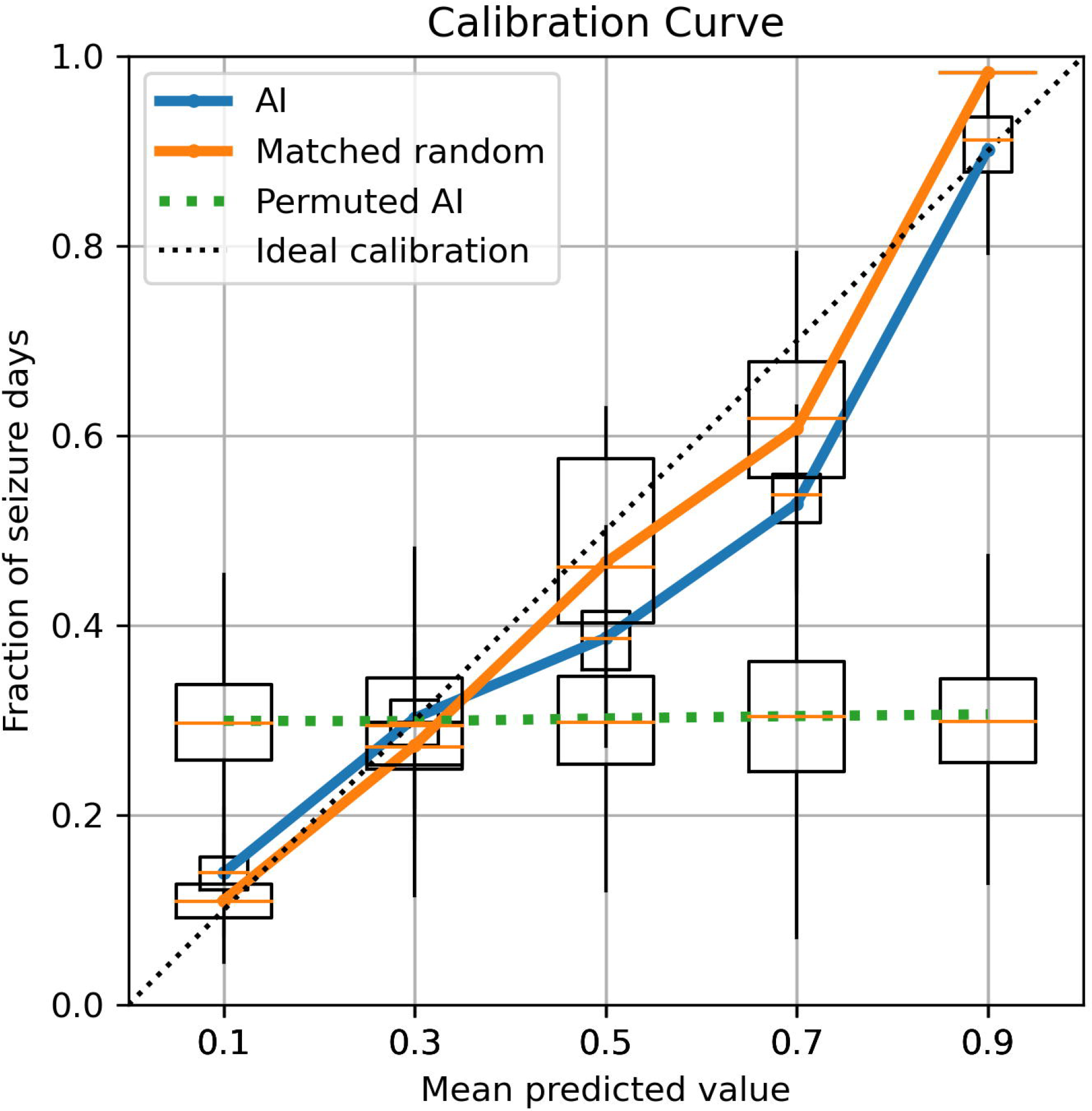
Calibration curves. The prospective seizure forecasts (pooled across all patients) are compared to the actual observed seizures for (1) the artificial intelligence (AI), (2) the rate matched random forecast (RMR), and (3) random permutations of the AI. Confidence intervals are shown by bootstrapping 1000 times (choosing patients with replacement). A perfectly calibrated (dashed line) forecast would always forecast the correct percentage of observed seizures. In this figure, the AI and random forecast deviate from the ideal somewhat, whereas the permuted reference is very poorly calibrated (as expected).

### Individual level results

In 7 patients (28%) the AI was superior (i.e., individual Brier Skill Score>0) to the random forecaster whereas for 9 patients (36%) the AI was superior to the moving average. The individual Brier Skill Scores (mean permuted AI forecasts^12^ as comparator) were median 0.00 (95% CI: −0.03 – 0.20). These values were notably lower than the group level BSS values (see Appendix I). Individual Brier Skill Scores with moving average as comparator were median −0.01 with 95% confidence range (−0.08-0.17). Individual level AI AUC values were very poor quality 0.43 +/- 0.21, as were individual level moving average values AUC 0.43 +/- 0.13.

Complete diaries with AI and moving average forecasts were plotted (Appendix D and E). There were 25 patients reporting less than 3 seizures in the pre-enrollment period (see Appendix D). Time-in-warning analysis was conducted (Appendix G).

The above analyses were also re-computed using the full set of 31 patients using the 3-month pre-enrollment diaries (Appendix F). This showed the AI was superior to random and moving average at the group level, and superior to the moving average at the individual level in 14 patients (45%). However, pre-enrollment data seizure rate was dramatically lower than the enrollment seizure rates, suggesting severe under-reporting.

The initial surveys (n=46), filled out prior to any forecasting, included questions related to seizure forecasting (Appendix G). Many (52%) patients stated they would not mind poor quality HIGH-RISK forecasts, and many (52%) did not mind poor quality LOW-RISK forecasts, yet almost all (91%) wanted access to forecasts. In the setting of LOW-RISK forecasts, 80% said they would not change their behavior, yet in HIGH RISK only 28% would not change – many stated that they would avoid risk-taking behavior (54%).

## Discussion

Our results prospectively attempted validation of a deep learning seizure forecasting system that is based entirely on seizure diaries. At the group level (considering all forecasts from all patients equally), one may mistakenly believe that the AI has strong potential. Using a random permutation surrogate as our comparator, the AI forecasts better than chance. However, a simple moving average forecaster turns out to perform just as well as the AI. Moreover, at the individual level (summarizing each patient separately first, then aggregating results), the AI outperforms the random permutation and the moving average in a small minority of cases, showing very poor overall individual level performance in AUC and Brier scores. The present work mirrors the previous retrospective study^8^, however it focuses on the individual patient level with physician curated, verified complete diaries. By reporting multiple metrics in different ways, this study highlights deficiencies of the present AI algorithm, and in certain outcome metrics. Clearly, the AI is not better than moving average forecasts; however, when missing data is present, the AI outperforms the moving average. Qualitatively, the data (Appendices D, E, F) suggests that at least one driver of periods of better forecastsrelates to the AI being better able to forecast multi-day clusters of seizures compared with the random permutation or the moving average. These clusters may reflect multi-day seizure susceptibility periods, though they do not appear to be periodic^3,13^, and they do not fit the classical definition of seizure clusters^14,15^.

Unlike our retrospective study^8^ that did not have verified complete diaries, the prospective study utilized weekly verified diaries from patients with clinical data confirming their epilepsy diagnosis. The misalignment of results between the former study and the present one may reflect the difference between the self-report and closely monitored self-report. In the case of the former, some events may be missed (under-reporting^16^), but in the case of the latter, some dubious events may be included (over-reporting^1^). There are no rigorous studies of over-reporting, which is challenging to accurately quantify. Here, the verified diaries have dramatically higher rates during the prospective phase compared to the pre-enrollment 3-month periods (see Appendix D) – strongly suggesting under-reporting.

The apparent under-reporting from the pre-enrollment period appears to reflect that without supervision, diaries might be incomplete. Our study required for enrollment the existence of a Seizuretracker account with at least 3-months of data prior to enrollment, however we did not verify or demand that such diaries were complete. This oversight is significant, because during the observed portion of the study we asked the participants weekly if their diaries were complete, and the seizure rates were consistently much higher (see Appendix D). Importantly, multiple lines of evidence^13,17–21^ show that, contrary to what we observed in our cohort, unverified seizure diaries often do reproduce patterns confirmed in verified systems, thus unsupervised seizure diaries may not always suffer from underreporting bias. Nevertheless, future studies will need to either confirm with participants that pre-enrollment diaries are complete or obtain longer duration observation periods and use only data obtained during confirmed timeframes.

Perhaps, one might suspect that patients with very high seizure rates would be unlikely to benefit from seizure forecasts at all. On the other hand, our cohort included only patients who wanted to be involved in a forecasting study (there was no compensation for this study), and 39% of them had very high seizure rates. Patient preferences (Appendix G) may even support inaccurate forecasts rather than no forecasts. It is worthwhile to note that the preferences reported were obtained prior to obtaining any forecasts from our team, therefore these can be viewed as the opinion of optimistic patients who had just enrolled in a study. Nevertheless, patients with less frequent seizure days are likely the most important to forecast (based on the need to make temporary changes in behavior), and the present algorithm did not excel in this area. More study is needed to better understand what the characteristics are of patients who would be most interested in seizure forecasts, and who would benefit most. It should be emphasized that in the absence of a nearly perfect forecast system, patients should never be encouraged to engage in risky behavior during periods of forecasted low risk.

The present study has several limitations. First, some people with epilepsy have very low (e.g., 1-2 seizures per year) or very high (i.e., ≥daily) seizure rates^22^. Such patients would not be likely to benefit from the current generation of daily forecasting tools. Second, it can be challenging for patients to maintain a seizure diary^23^, thus limiting tools of this nature to patients and caregivers willing to maintain a diary. Third, our prior^8^ and present study did not have available EEG data to augment forecasts. Although speculative, including EEG data may enhance the performance of these models. Fourth, the 5-month prospective duration of the present study may be too short to make definitive conclusions about the utility of the AI algorithm. To address this deficiency, our group will be conducting a larger study soon with a longer observation period to allow for sufficiently large windows of investigator-verified seizure diaries. Sixth, there was a presumed dramatic under-reporting in the pre-enrollment period. In our future study, we will not include a pre-enrollment period due to the challenges in verifying that they are complete. Finally, the choice of reference standard comes at a cost. Our average permutation (a.k.a. random) forecaster standard could not be realistically provided to patients in real-time. Conversely, our second reference standard was the moving average forecaster. This can be implemented in a real-time system, making it a realistic comparator A comparison of the calibration curve (Figure 1) shows very poor calibration of the permuted AI, but decent calibration of moving average and AI. In using both, we highlight the advantages and disadvantages of each.

We hope that future advances in wearables^6^ and minimally invasive tools^7,24^ can synergistically be applied to diary-based forecasting tools to achieve higher accuracy and wider patient appeal.

## Data Availability

Data is available on reasonable request.

https://github.com/GoldenholzLab/deepManCode

## Data availability

Data is available on reasonable request.

## Epilepsia ethical publication statement

We confirm that we have read the Journal’s position on issues involved in ethical publication and affirm that this report is consistent with those guidelines.

## Acknowledgements and Funding

DMG was supported by NINDS KL2TR002542 and K23NS124656. MBW received funding support from the American Academy of Sleep Medicine through an AASM Foundation Strategic Research Award; the NIH (R01NS102190, R01NS102574, R01NS107291, RF1AG064312, RF1NS120947, R01AG073410), and NSF (2014431). Dr. Westover is a co-founder of Beacon Biosignals, and Director for Data Science for the McCance Center for Brain Health.

## Author contributions

DG, MW, and RM contributed to conception and design of the study; CE, DG and RM contributed to acquisition and analysis of data; DG drafted a significant portion of the manuscript and figures.

