## Appendix for "Prospective validation of a seizure diary forecasting falls short"

A. **The AI model and the Moving Average forecast**

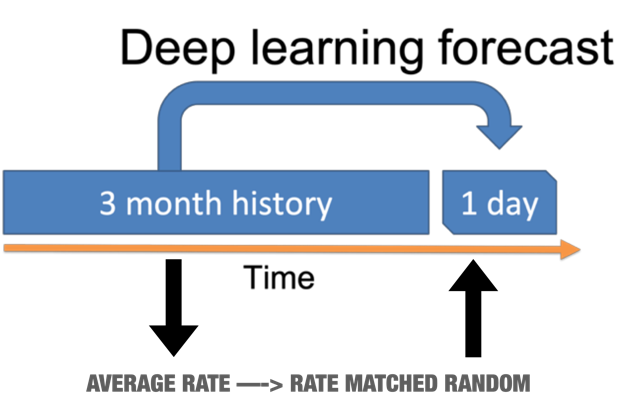
The AI model was developed in a previous study^1^ using data from 3806 patients for training. In the training set, model hyperparameters were established and fixed. The input to the model was 84 whole numbers, representing the number of seizures in each of 84 days leading up to the forecasted day. The model itself was implemented in python using Keras and TensorFlow, with 2 LSTM layers connected to 3 fully connected layers (recurrent neural network connected to a multilayered perceptron). In the original study, 2 additional input streams were considered as a hyperparameter, one for average seizure duration per day, and one to flag if each seizure was a generalized seizure. This additional input was found to be not needed in the final model based on the training data and was excluded from further testing on the testing set in the original study.

The moving average forecast simply used the average number of seizure days in the 3-month history to predict the risk of a seizure in the coming 24 hours. Let each historical i^th^ day be represented by d_i_, which is a dichotomous variable indicating seizure day (=1) or non-seizure day (=0). The forecast is given from the 84-day moving window:

$moving average= \frac{1}{84}\sum_{i=1}^{84} d_{i}$	(Eq. 1)

It is important to note that the moving average forecast value ranges between 0 and 1, by definition, thus crudely approximating a “probability”. The moving average model is a modification of our previous “rate matched random” model, however the AI model is unchanged from the previous study.

The AI model was used “as is” from the prior study, including all the previous training weights, to properly validate the new prospective data while avoiding data leakage.

The full code is available here: <https://github.com/GoldenholzLab/deepManCode>.

B. **Brier Skill Score**

The Brier Skill Score is used to compare two forecasting algorithms. A Brier Skill Score of 1 represents the candidate algorithm is perfect, 0 indicates a candidate is not better than the alternative, and -1 indicates the alternative is perfect. The basis of the score is a ratio based on Brier Scores^2^. The Brier score is commonly used to measure the accuracy in weather forecasting tools and is calculated as follows. Let N be the total number of observations, with the i^th^ dichotomous observation denoted o_i_, and the forecasted value (a probability between 0 and 1) for the i^th^ observation denoted f_i_:

$Brier score= \frac{1}{N}\sum_{i=1}^{N} {(f_{i}-o_{i})}^{2}$	(Eq. 2)

Brier scores range between 0 and 1, with scores closer to 0 being more accurate, and closer to 1 being more inaccurate. Let the Brier Score of algorithm C (the candidate model) be B_C_, and the Brier Score of algorithm R (the reference model) be B_REF_. The Brier Skill Score is given by:

$Brier Skill Score=1-\frac{B_{C}}{B_{REF}}$	(Eq. 3)

The Brier Skill Score takes on values between 1 and -1. A value of 1 would indicate a perfect candidate, a value of >0 indicates the candidate represents an improvement over the reference, and <0 indicates that the reference is superior to the candidate. Therefore, BSS should be >0 for any forecasting tool compared to a reference standard such as the random algorithm described in Appendix A.

To compute individual level BSS values, each patient was considered separately. A BSS was computed (Eq 3) for each patient. The median and 95% confidence interval values across patients were reported.

To compute group level BSS values for M patients, the entire collection of forecasts from all M patients were concatenated into a single set of f_i_ values, and the corresponding true outcomes were concatenated into a single set of o_i_, values. The computation of Brier Score (Eq. 2) and Brier Skill Score (Eq. 3) were used with these larger sets. By bootstrapping with replacement of patient, a median and 95% confidence value was obtained for each group level BSS.

C. **Using as much data as possible**

The AI required 84-day history to produce a forecast of the risk for seizure in the subsequent 24-hour period. The 84-day history could come from either a) the 3-month pre-enrollment phase, b) from the prospective period, or both. The forecasted days only came from the prospective period, during which the diaries were verified to be complete. In some cases, participants did not verify every week. This produced “holes” in their diary. Similarly, some participants dropped out early. In both cases, we attempted to maximize the number of forecasts possible from each participant. For example, if a participant dropped out after 16 weeks but verified every day of those 16 weeks as complete, then we included 16 weeks of forecasts. For another example, if a participant did not verify the 17^th^ week, but every other week before and after were verified, we did the same analysis – including the first 16 weeks only. Conversely, if a participant verified every week except the first week, then the pre-enrollment 84-day history was not used, and the first 84 days of verified history were used to make a forecast on the 85^th^ (and subsequent) days. The grid is provided below for all participants that initiated a connection to the Seizuretracker.com database to show how many of each situation is represented. Start and end dates were removed to protect patient privacy.

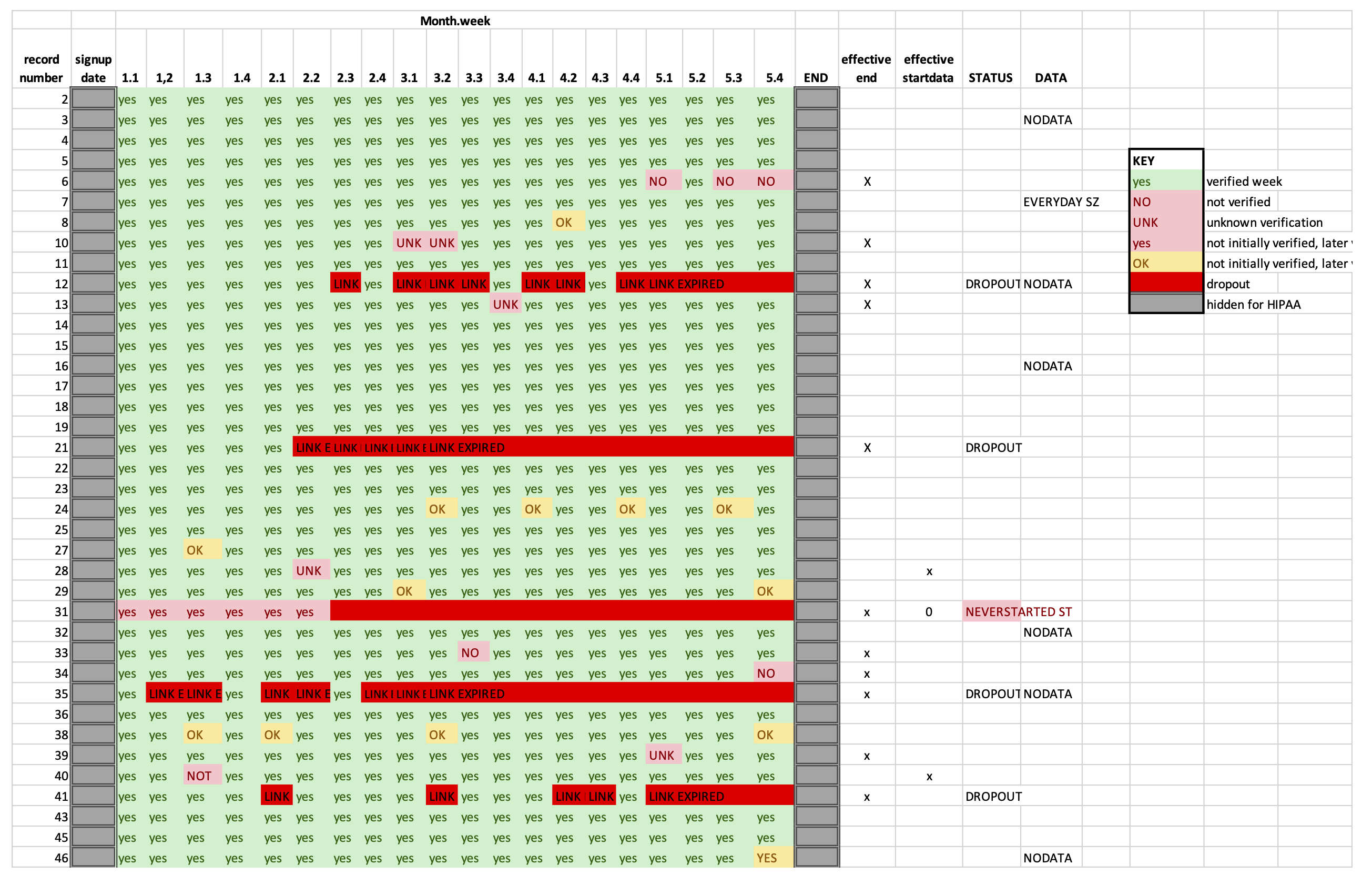

D. **Example forecasting patient with 3-month pre-enrollment**

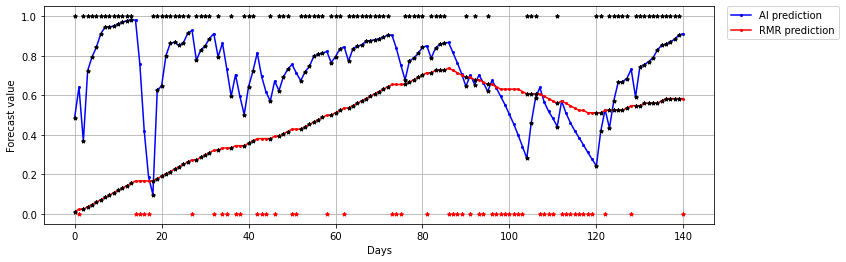
An example patient is shown here with black dots for seizure days shown at the y=1.0 position, and red dots for non-seizure days in the y=0.0 position. The moving average forecast is labeled “RMR” and plotted alongside the AI forecast. As can be seen in this example patient with a very high seizure frequency, there forecasts are not periodic, and they appear to incorporate a “memory” of recent seizures with more weight on very recent events. The risk appears to increase after a recent seizure and decrease after a lack of recent seizure, but there is an accumulated risk from previous days that is apparently factored in. The rate matched random forecaster is much smoother, yet it provides a much less accurate forecast.

We note that the RMR forecast appears to ramp up during the initial half of the record above. This reflects how many seizures were recorded in the 84-day history *prior to* the beginning of the prospective period. All subjects stated that they maintained an accurate diary that included data 3 months prior to the prospective study beginning. That said, examples like this one suggest that many patients may have reported fewer seizures during the 3 months prior to the prospective study start date. Patients in our study were reminded weekly to complete their seizure diaries and were asked if each week included all events. The 84 days prior to the start of the study did not have these reminders. Many patients appeared to report a lower event rate during the 3 months prior to the start of the prospective period compared to their 5 month “typical rate”. It is likely this reflects under-reporting, or reflect fluctuations in event rates. In the absence of objective data (such as implanted EEG), it would be impossible to be sure what is the true cause of this difference. Nevertheless, what can be seen in the example patient is that the RMR under-performs particularly prior to the acquisition of *prospective* 84 days in the moving window. The AI performed better throughout, possibly because less weight was attributed to events further in the past.

E. **Individual forecast plots including 3-month pre-enrollment:**

The results of analysis when the 3-month pre-enrollment data is included comes to a different set of conclusions (which we believe are erroneous):

Of 46 recruited participants, 1 was ineligible, 3 were seizure-free, and 11 provided insufficient diary data. Within the remaining 31, there were 3 dropouts, and 8 who missed some of the weekly diary completeness responses. Forecastable diary days (Appendix C) ranged 15-141 (median 141) days. Total seizures per patient ranged from 1-140, (median 27). Participant characteristics are summarized in Table 1 in the main manuscript.

The AUC across all 31 patients was AI: 0.82 [95% CI 0.74-0.88] and moving average: 0.78 [95% CI 0.70-0.84]. The AI AUC was higher than moving average AUC (Mann-Whitney U, p<0.0001). The Brier score was AI: 0.15 and moving average: 0.22. The overall Brier Skill Score (AI vs moving average) was 0.13 [95% CI 0.02-0.25] (Figure 1). In 13 patients (42%) the AI was superior to moving average (i.e., individual Brier Skill Score>0). Individual Brier Skill Scores were median -0.03 with 25%-75% range (-0.07-0.10). Complete diaries with AI and moving average forecasts were plotted (Appendix D and E).

Brier Skill Scores comparing Individual AI forecasts to mean permuted AI forecasts showed median -0.01 (25%-75% range -0.03 – 0.04). There were 14 patients with Brier Skill Scores>0, i.e., better than chance.

Shown here are the complete forecast plots for all non-degenerate patient diaries (degenerate meaning diaries listing all days with seizures or no days with seizures). Time is represented on the horizontal axis, and after time 0 is when forecasts begin. The initial 84 days leading up to day 1 are shown without forecasts, because these are used to produce the first forecasted day. Seen clearly here, many patients reported zero or very few seizures in their 84 days leading up to day 1. This strongly suggests under-reporting during the 3 months prior to the curated portion of the study, which has the effect of diminishing the effectiveness of the random forecast until sufficient “typical” seizures are reported.

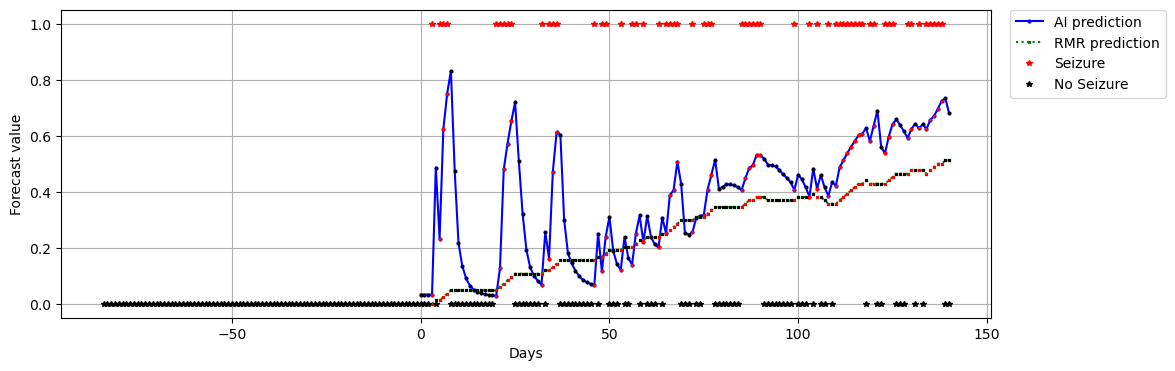

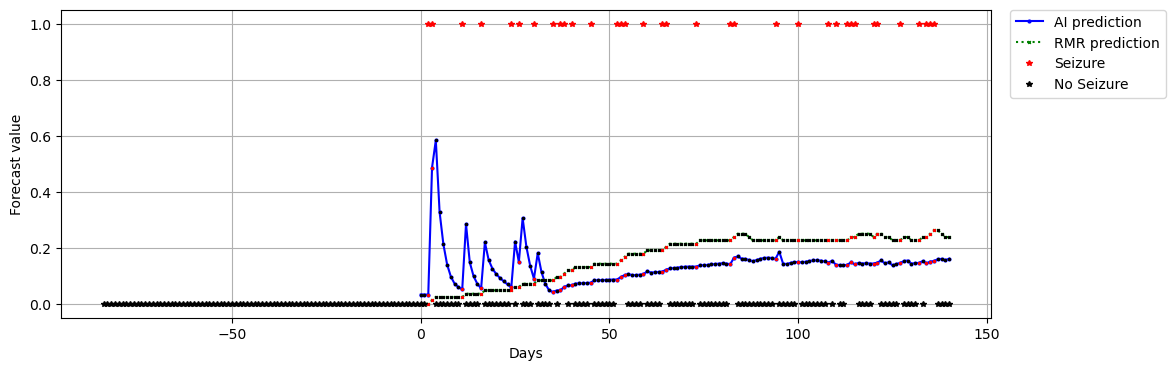

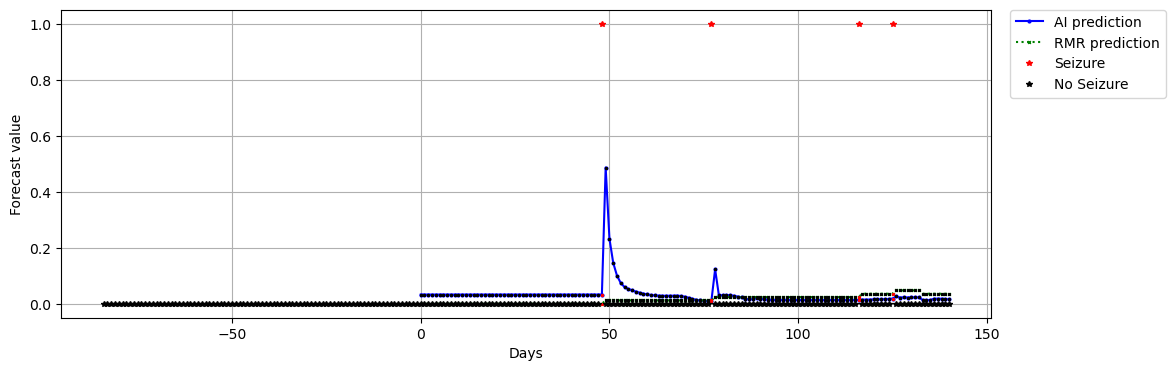

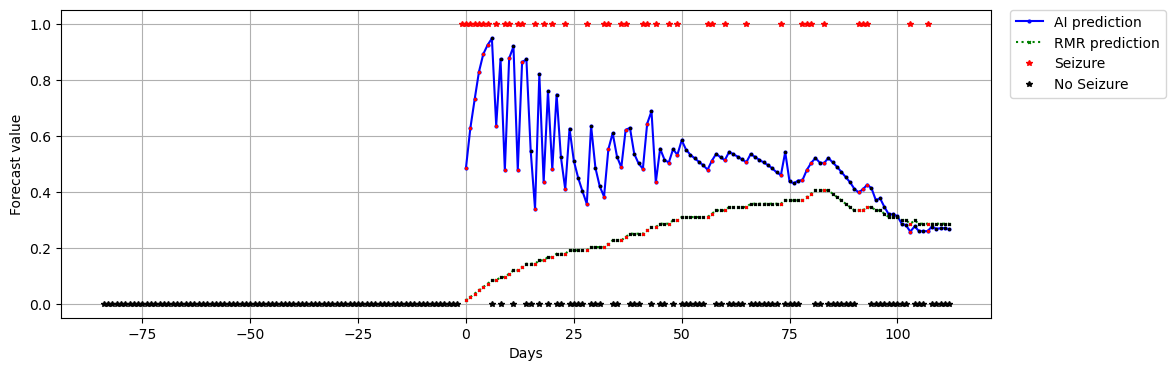

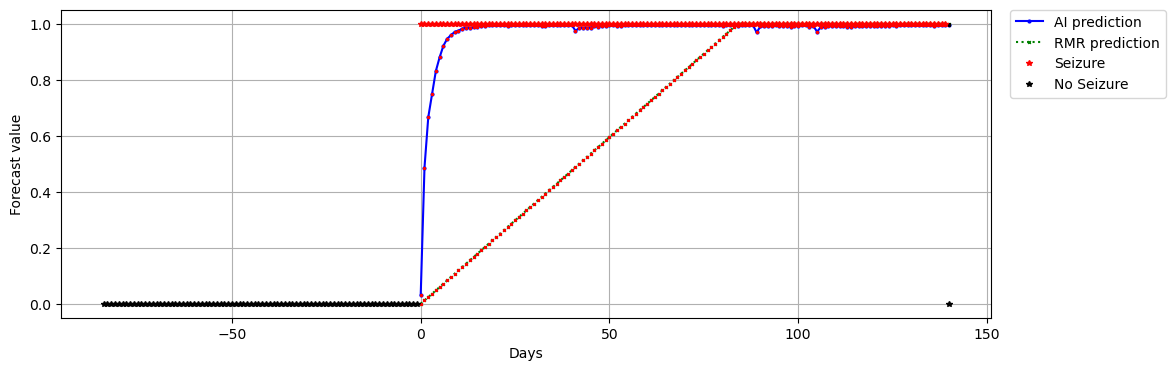

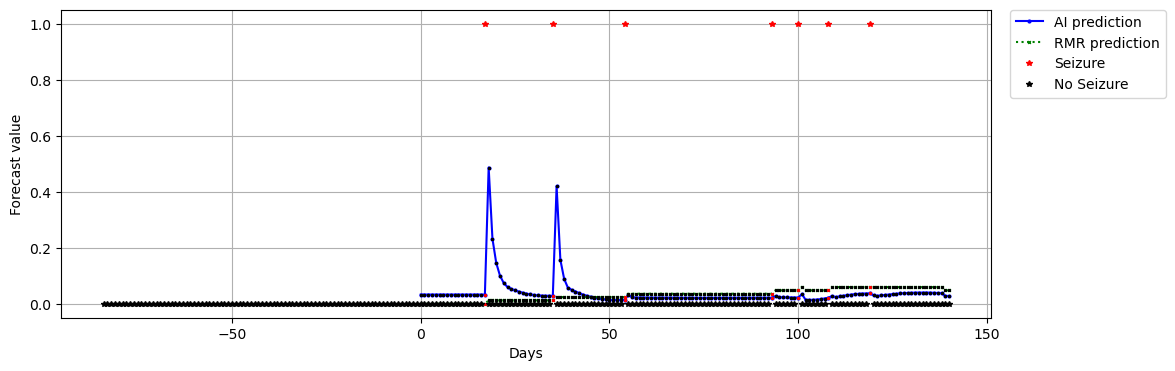

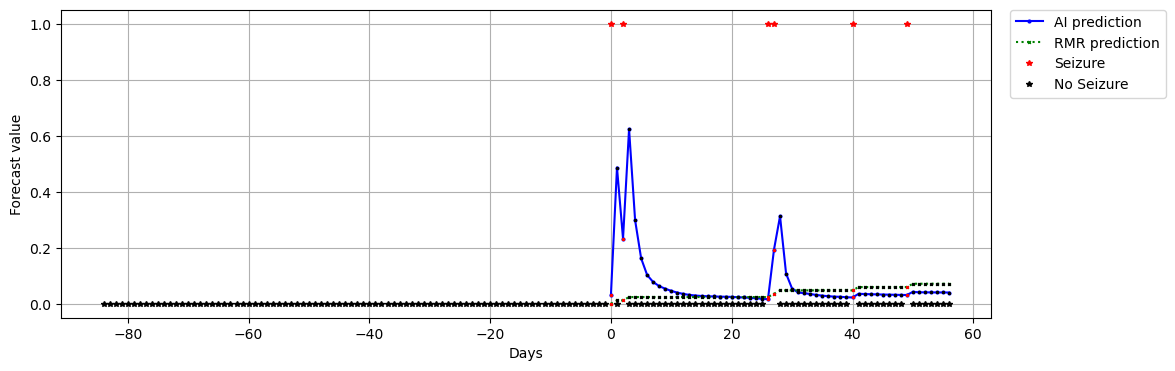

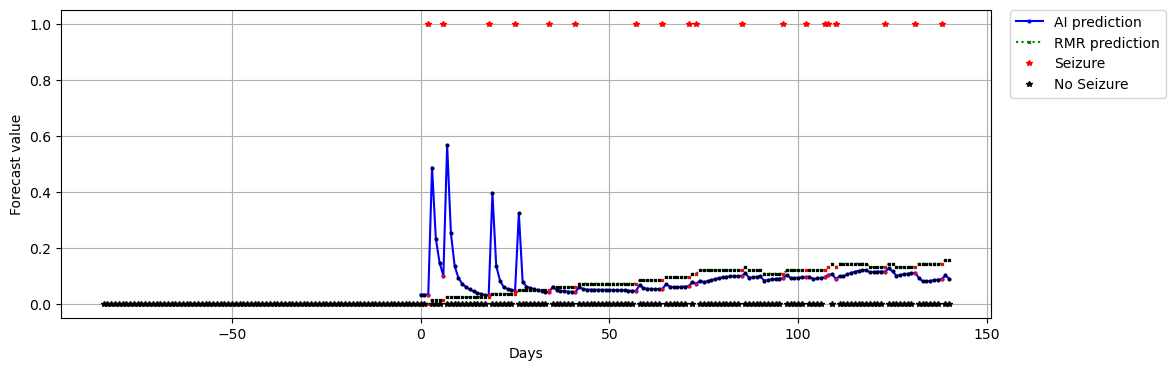

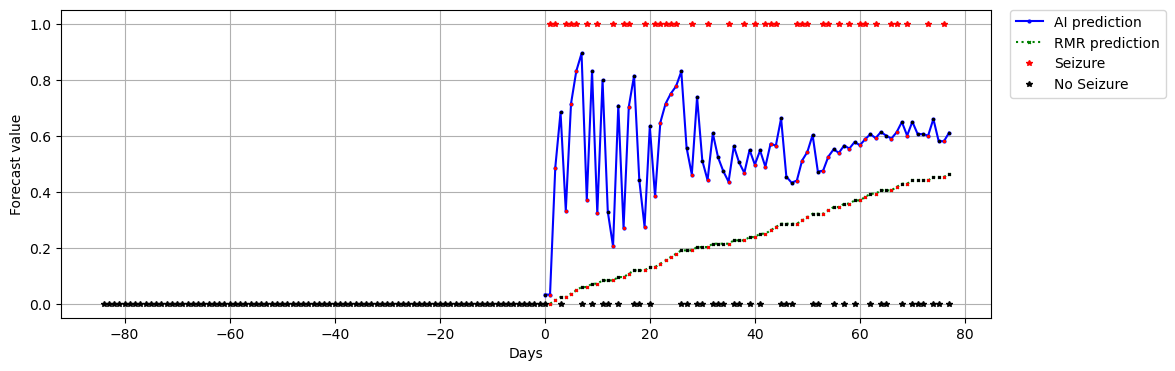

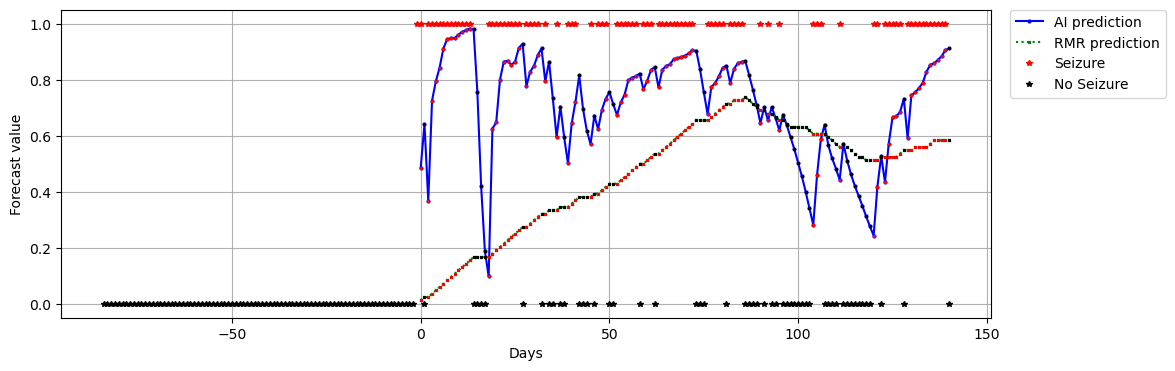

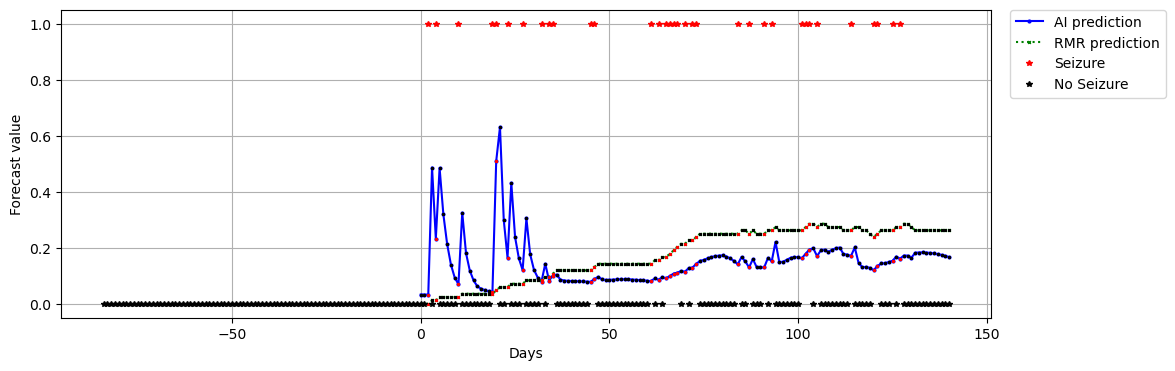

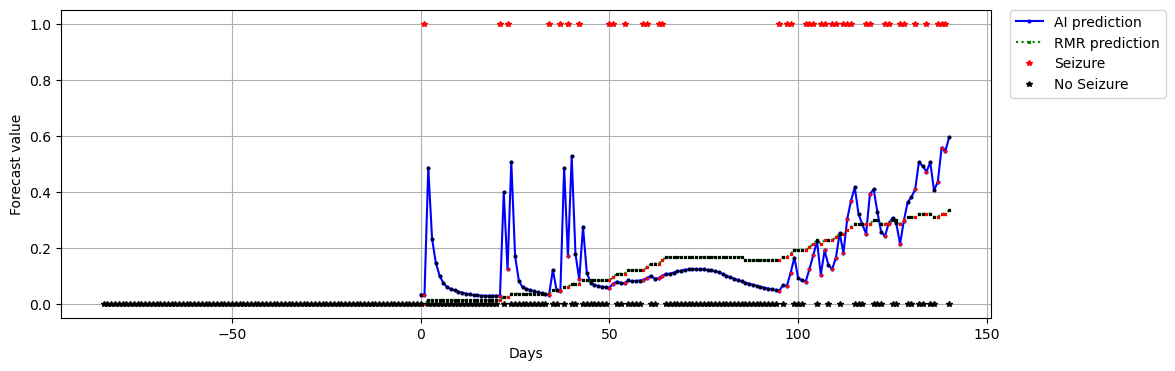

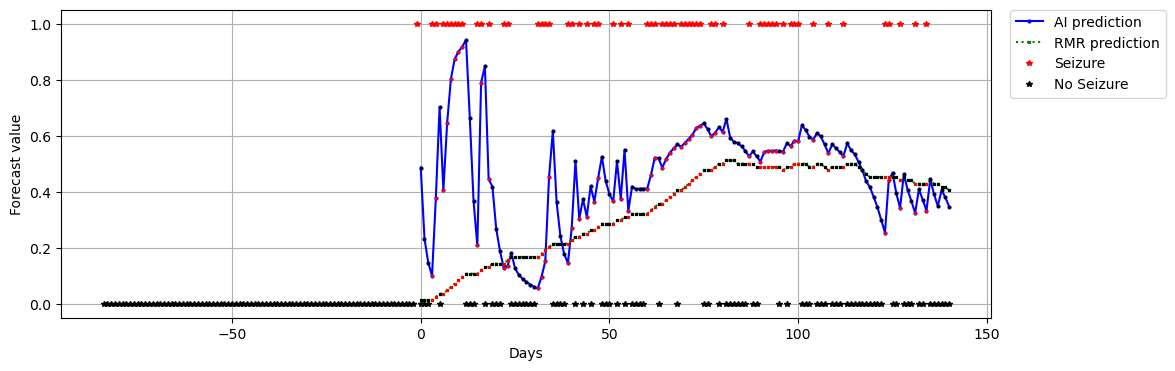

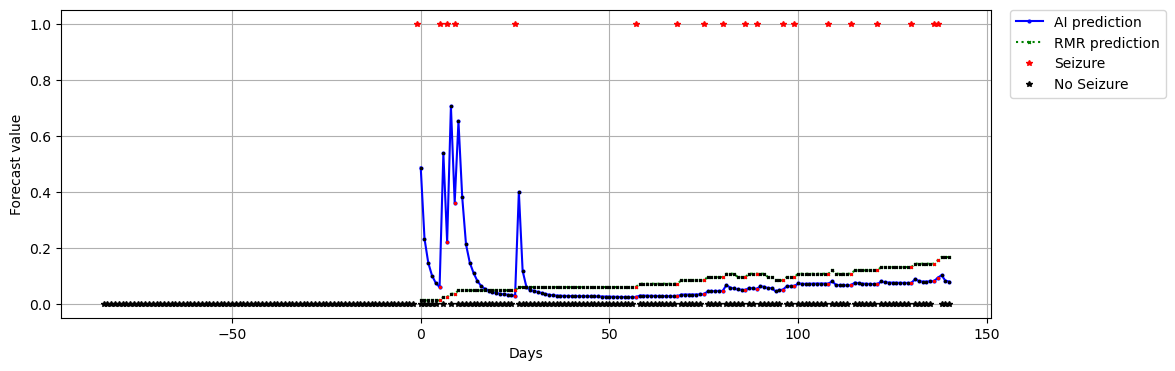

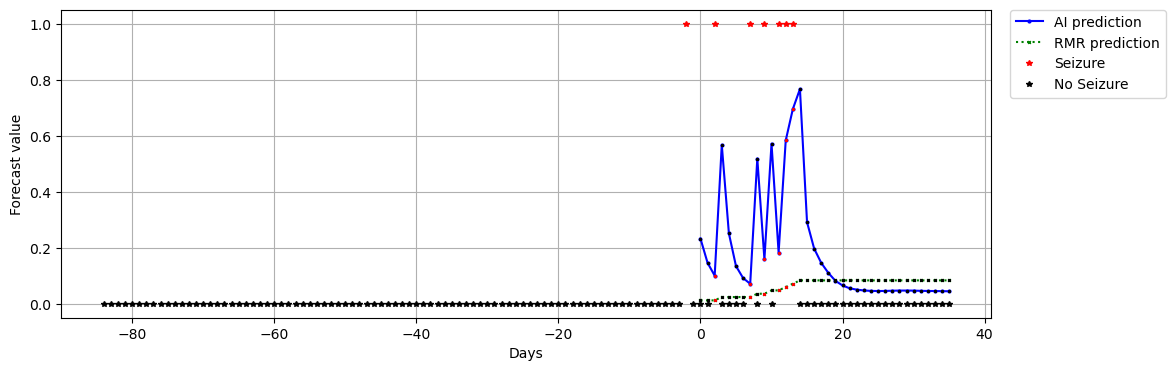

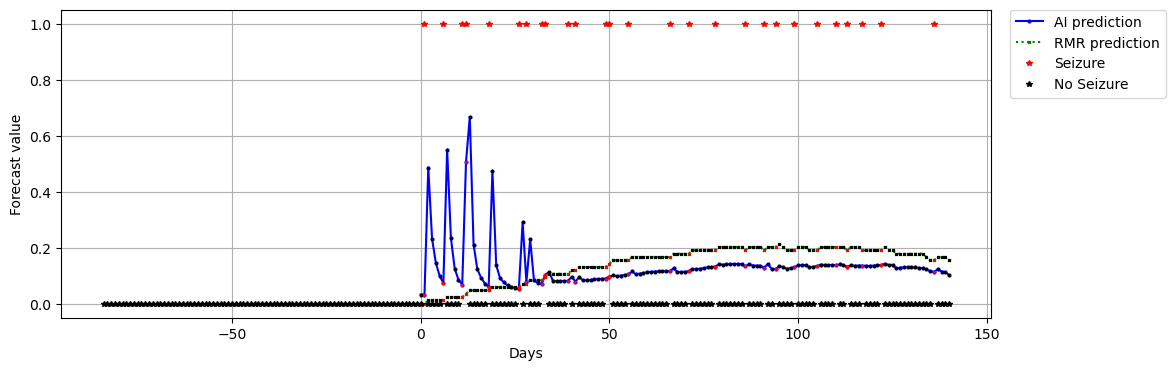

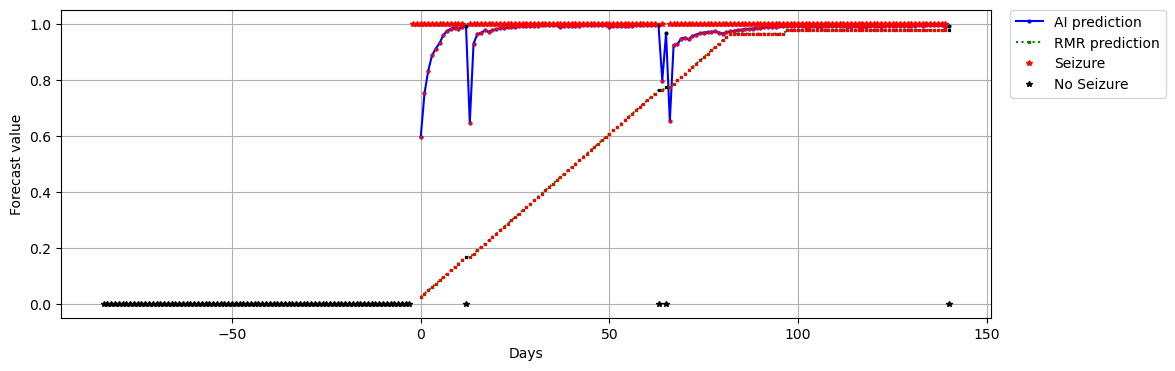

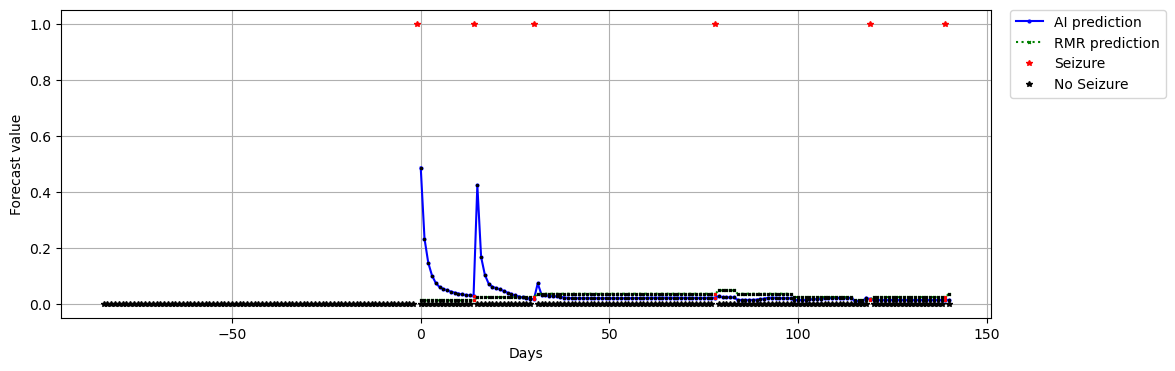

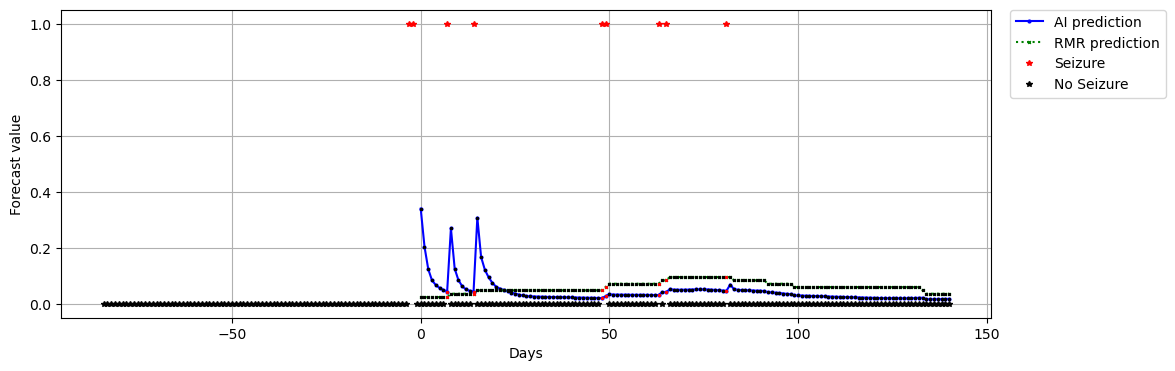

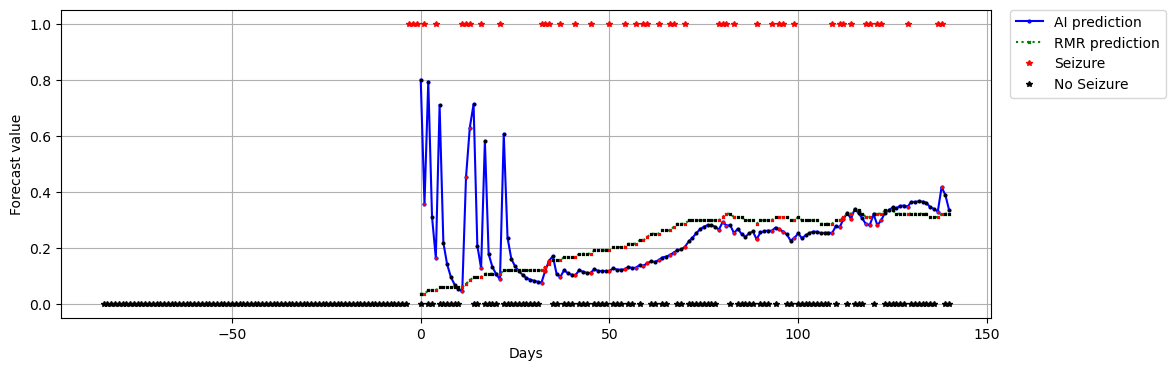

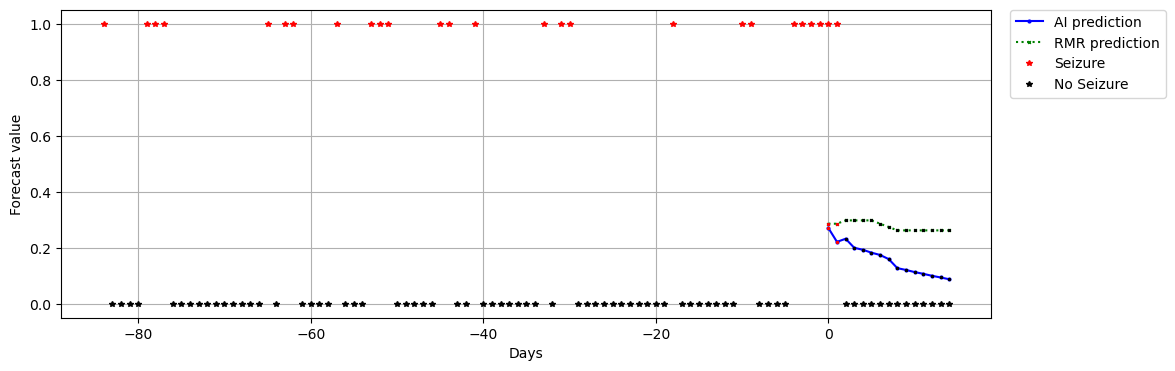

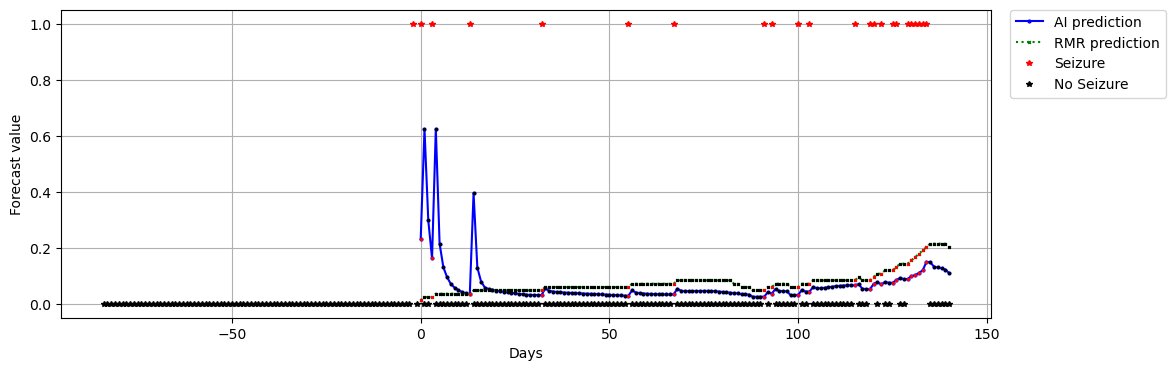

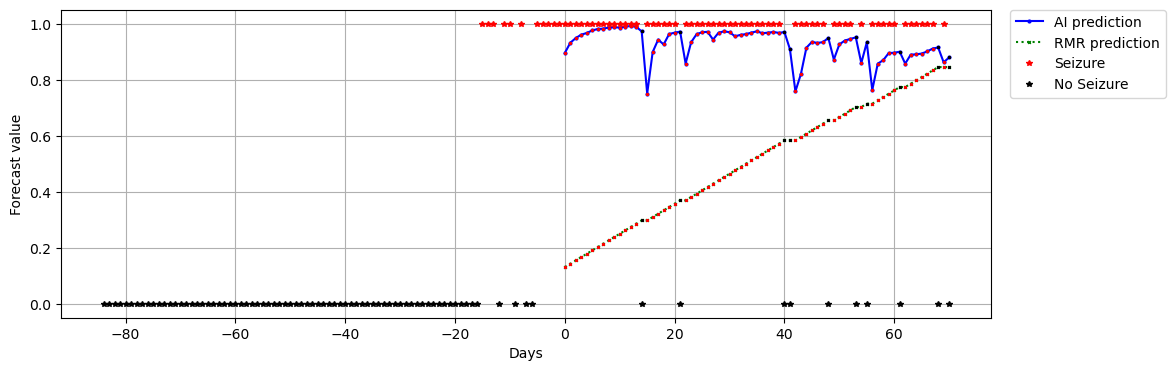

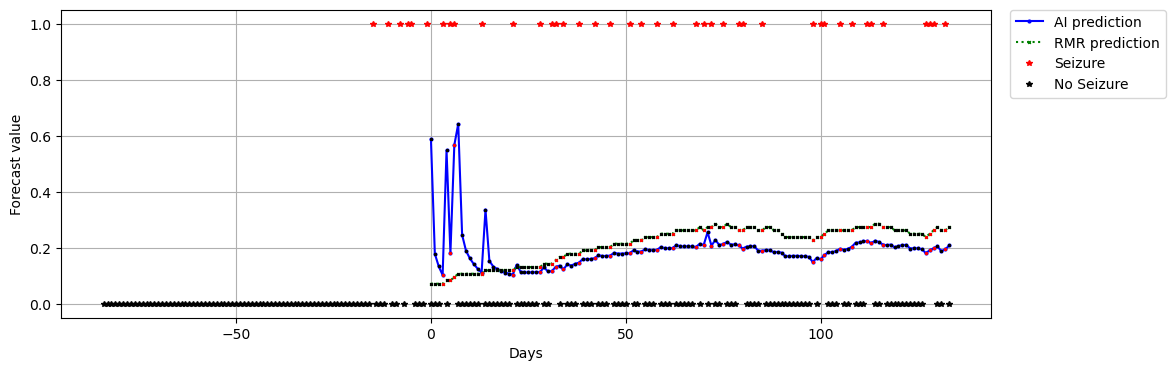

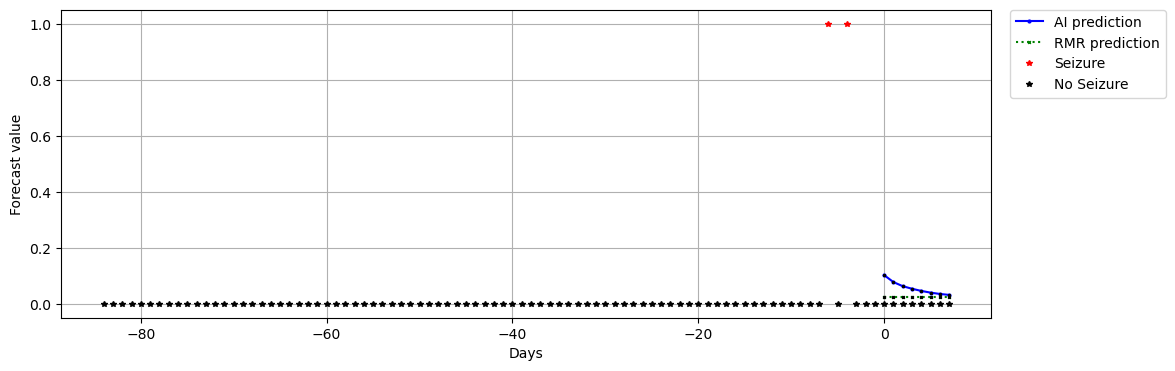

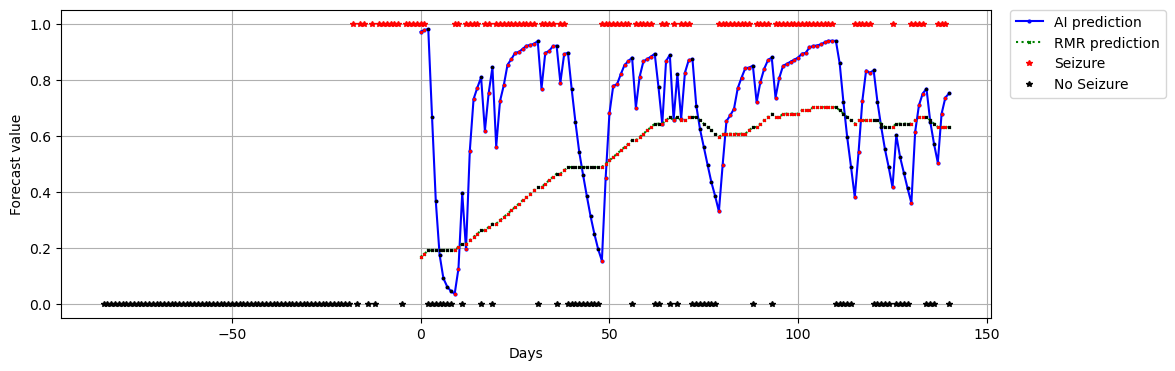

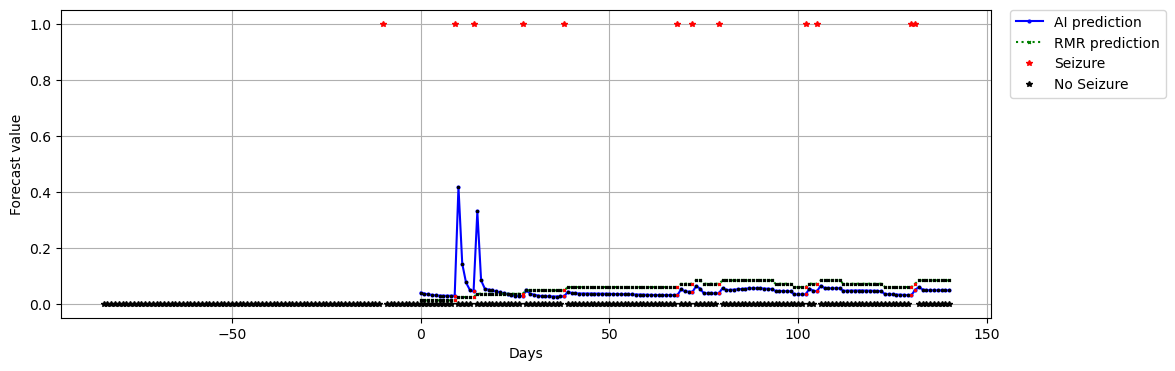

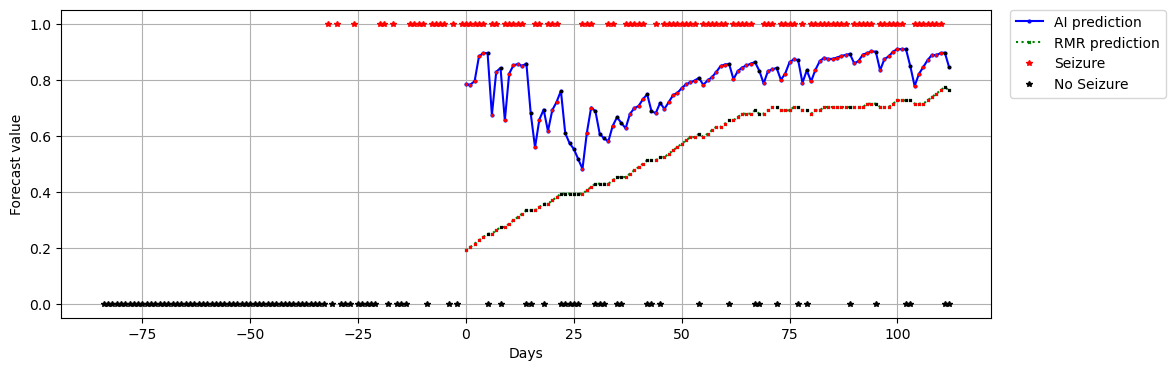

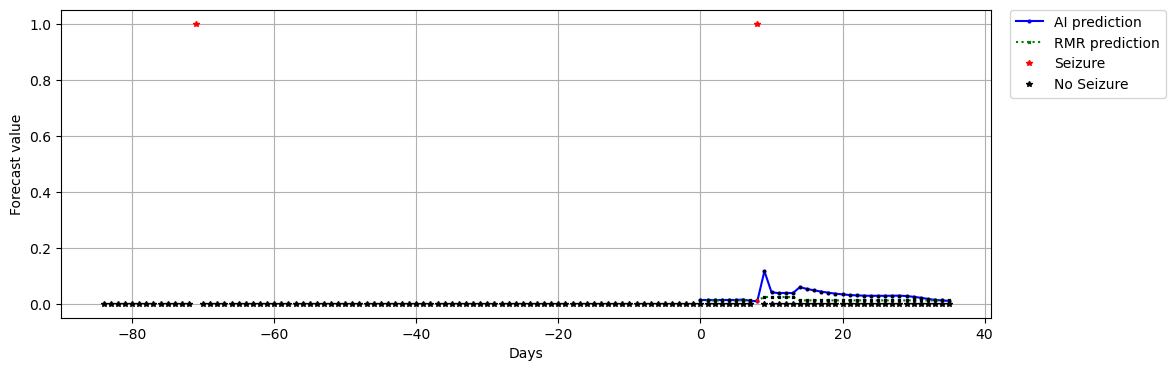

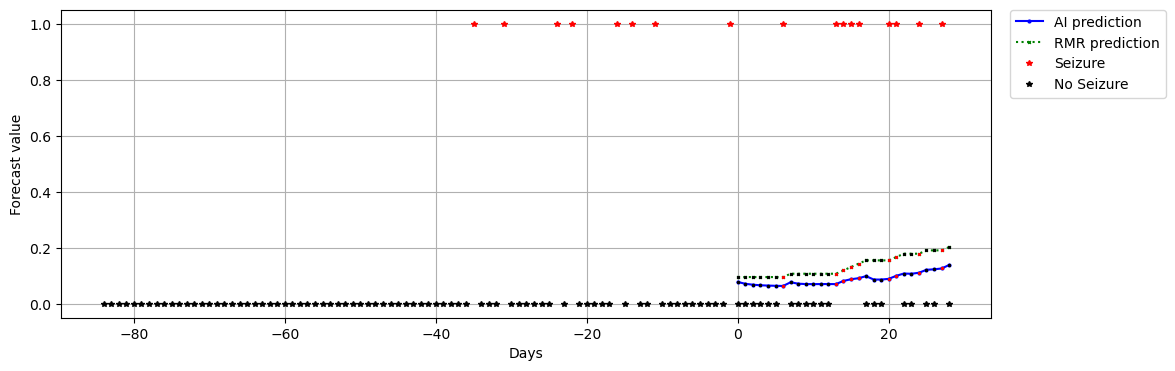

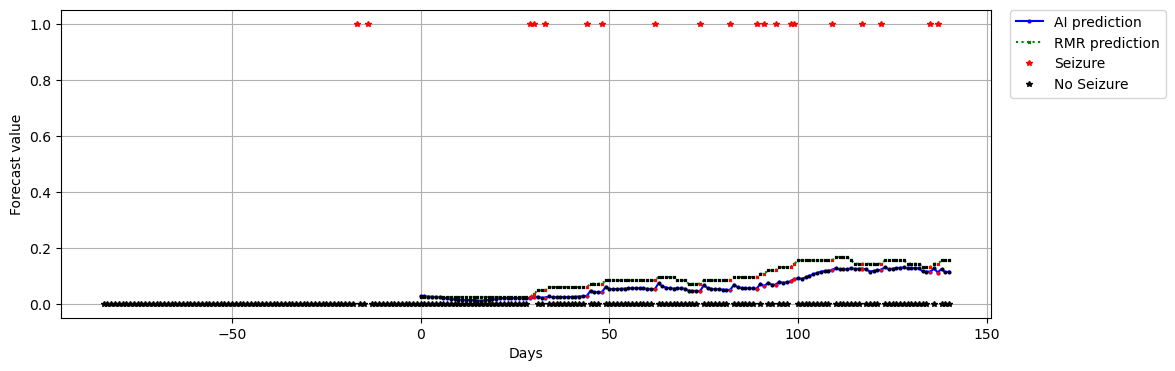

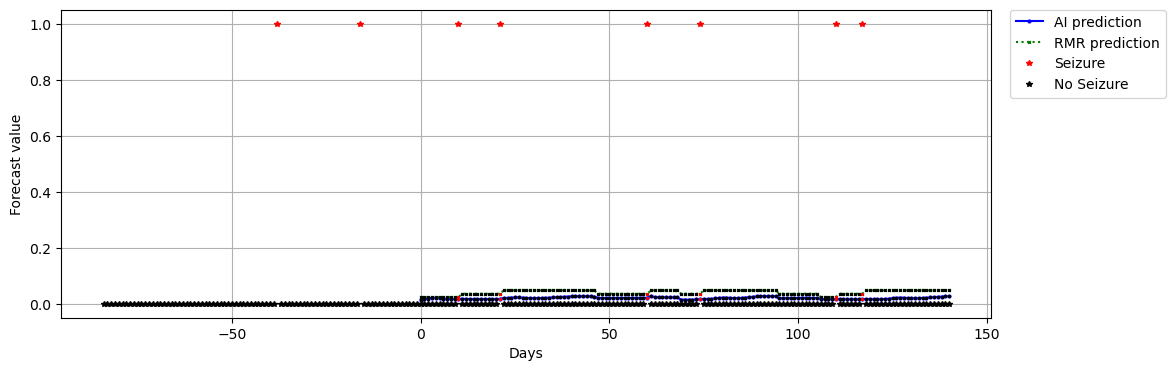

F. **Complete diaries, excluding 3-month pre-enrollment data:**

Shown here are the complete forecast plots for all non-degenerate patient diaries (degenerate meaning diaries listing all days with seizures or no days with seizures). Time is represented on the horizontal axis, and after time 0 is when forecasts begin. The initial 84 days leading up to day 1 are shown without forecasts, because these are used to produce the first forecasted day. Seen clearly here, many patients reported zero or very few seizures in their 84 days leading up to day 1. Here, we do not include the 3 month pre-enrollment period, given the possibility of under-reporting during that time.

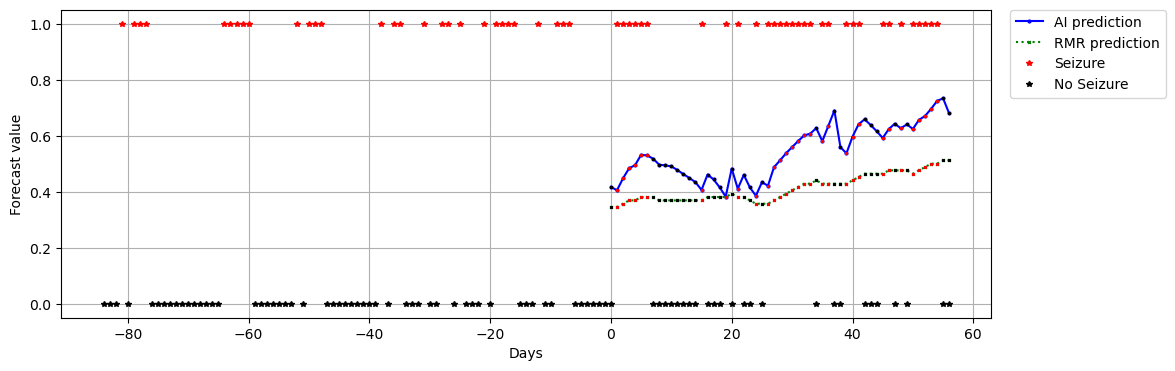

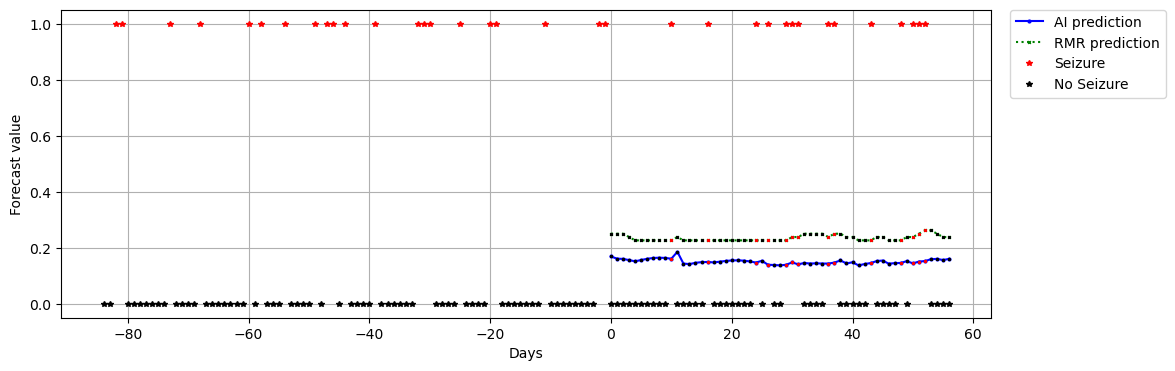

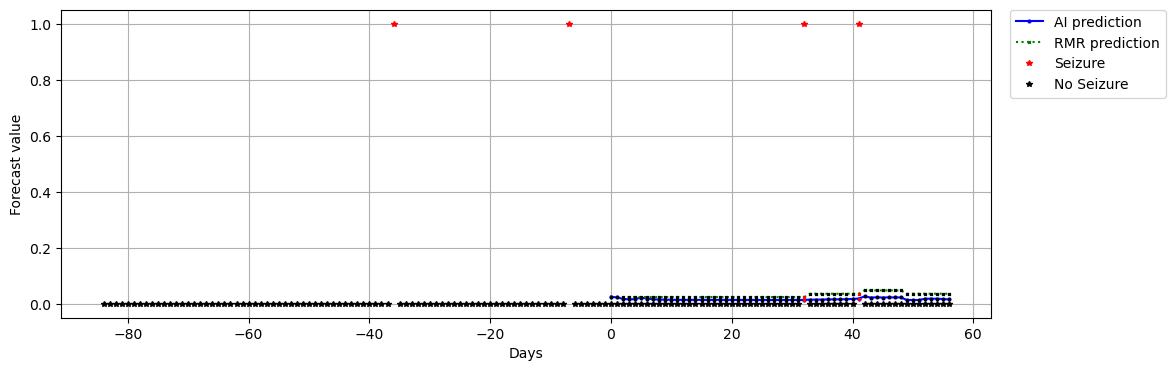

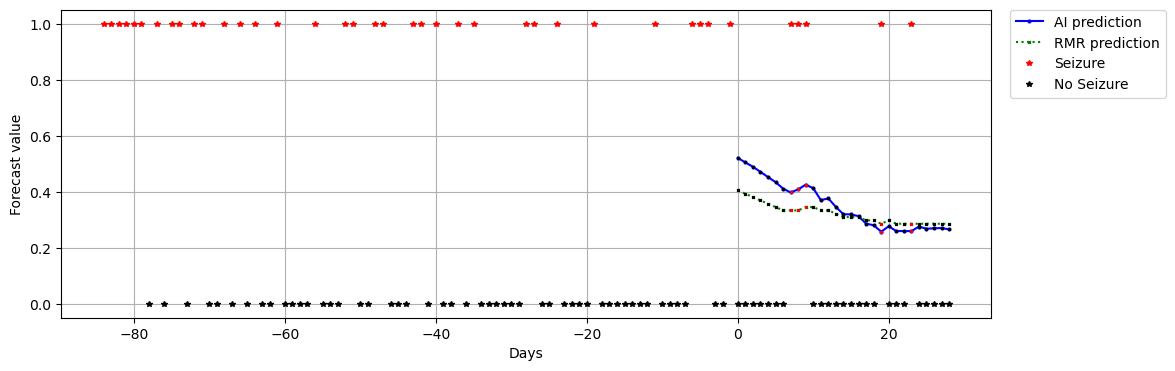

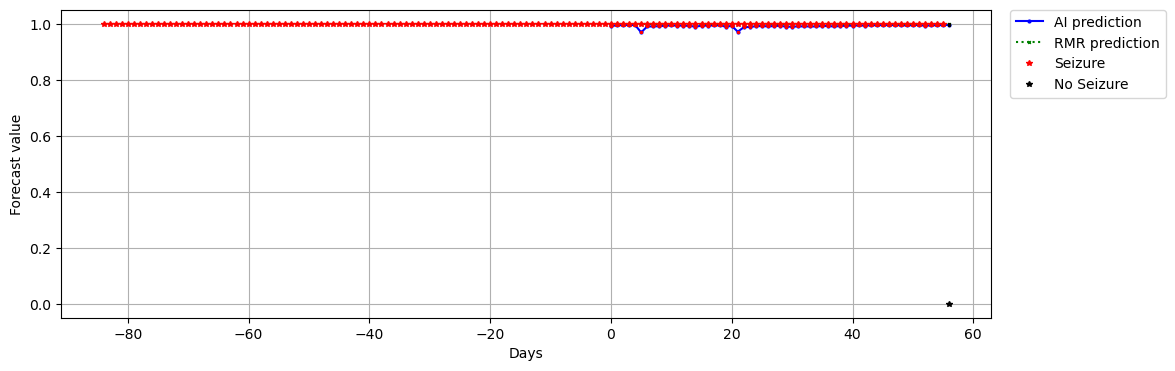

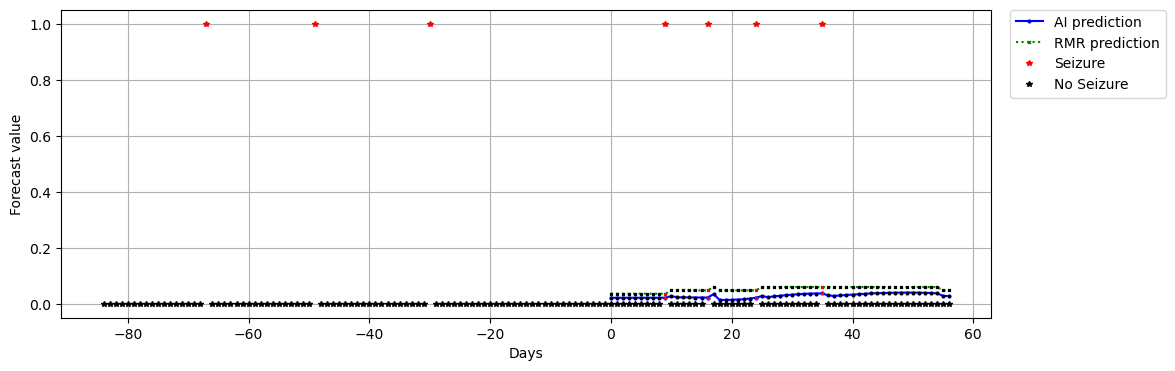

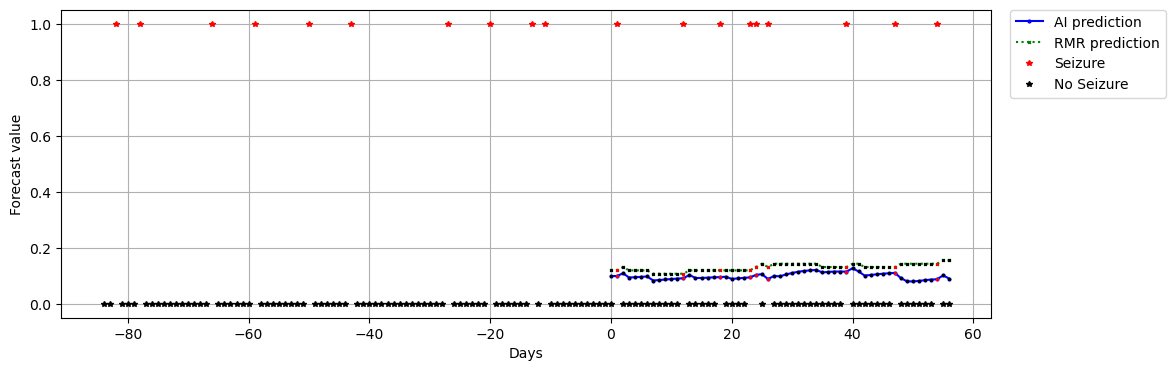

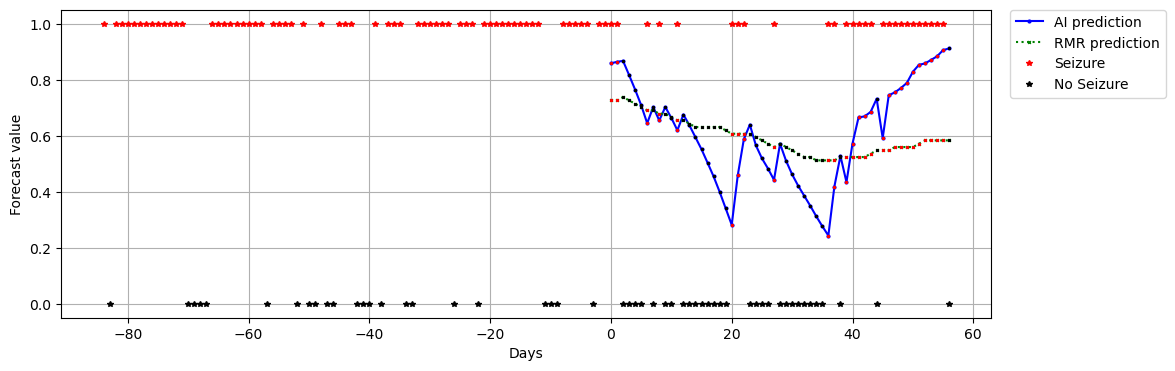

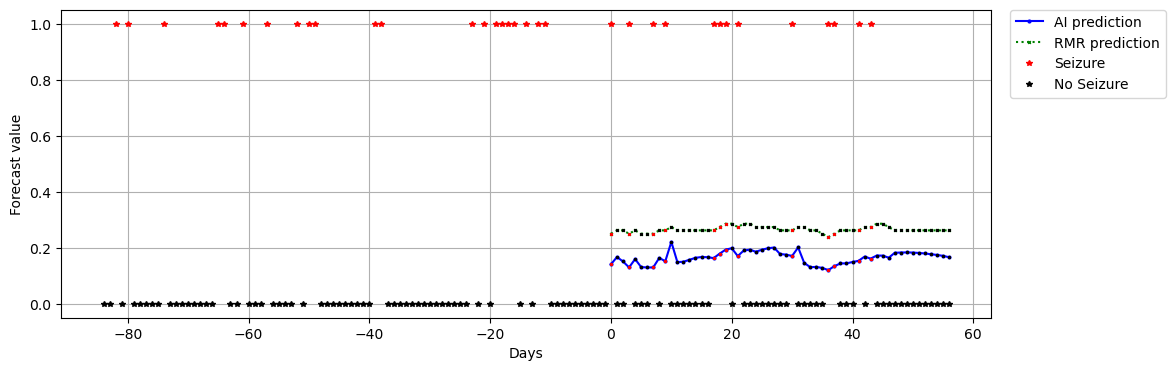

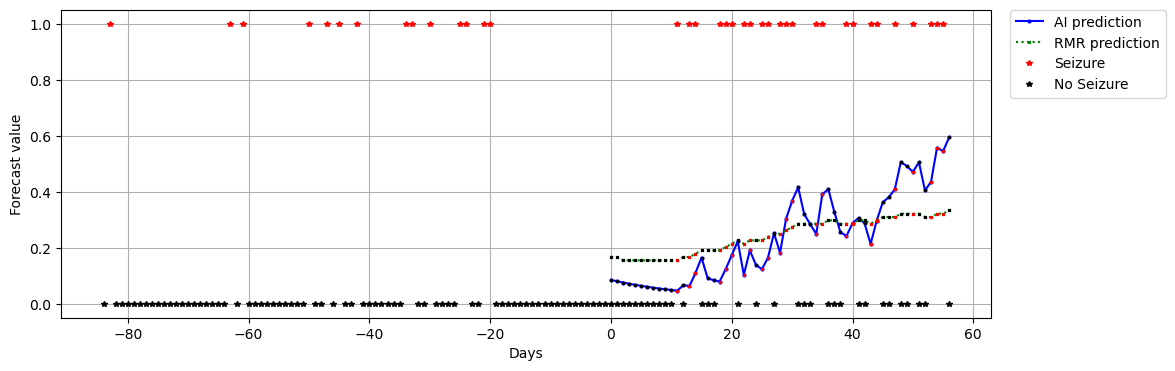

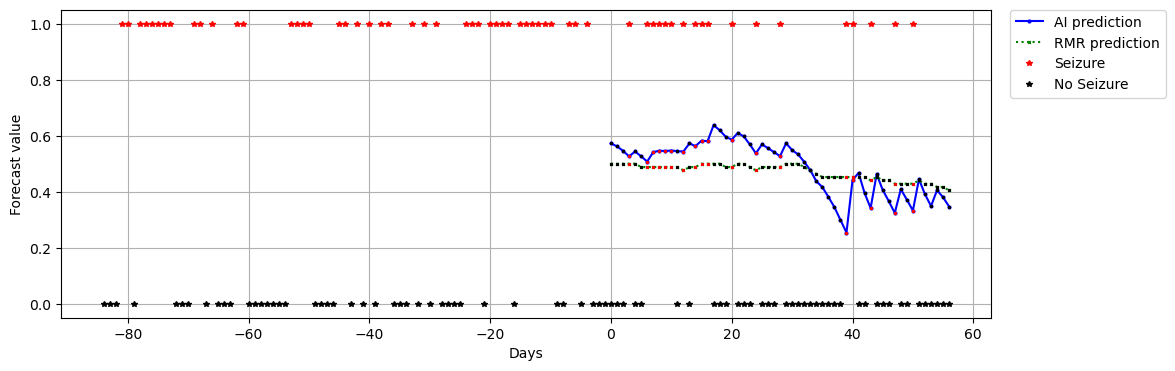

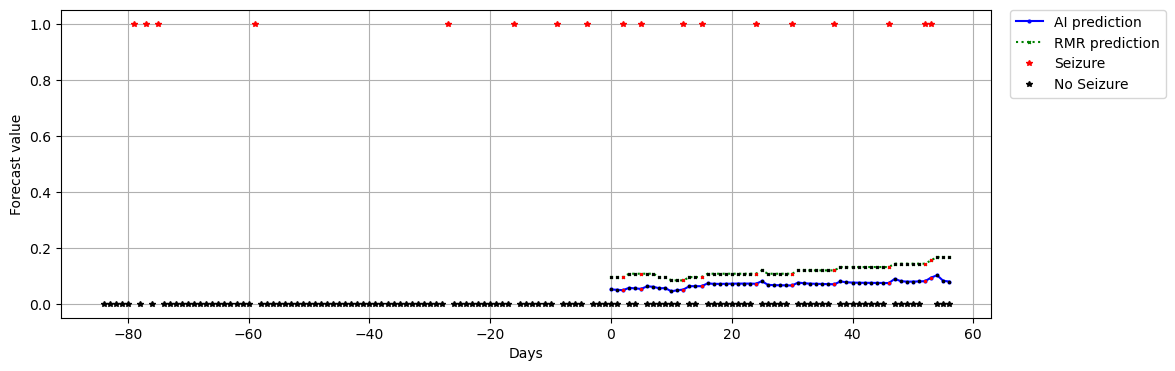

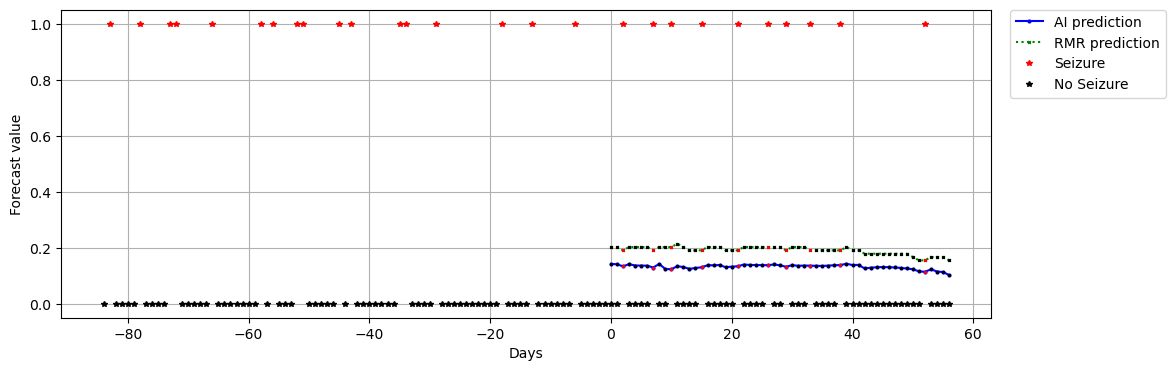

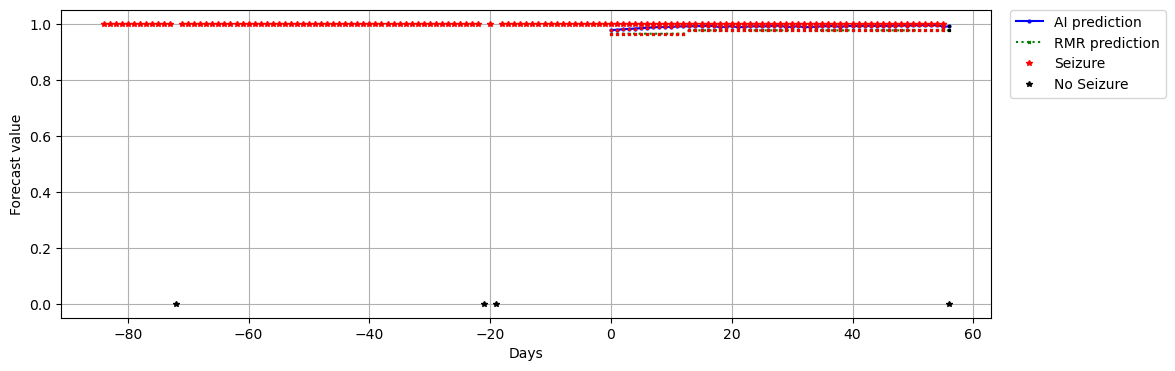

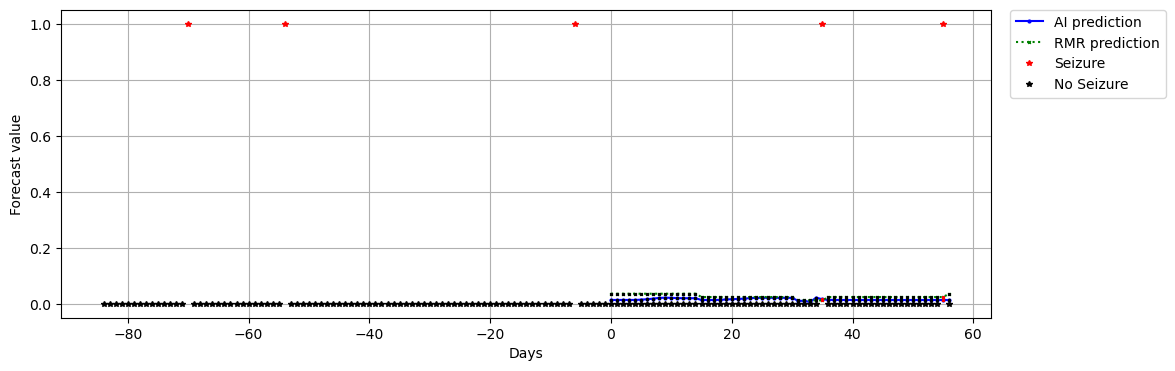

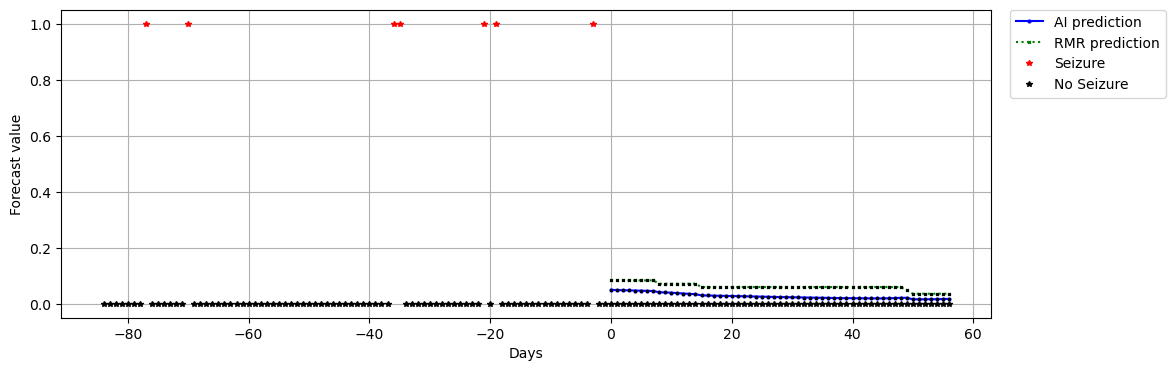

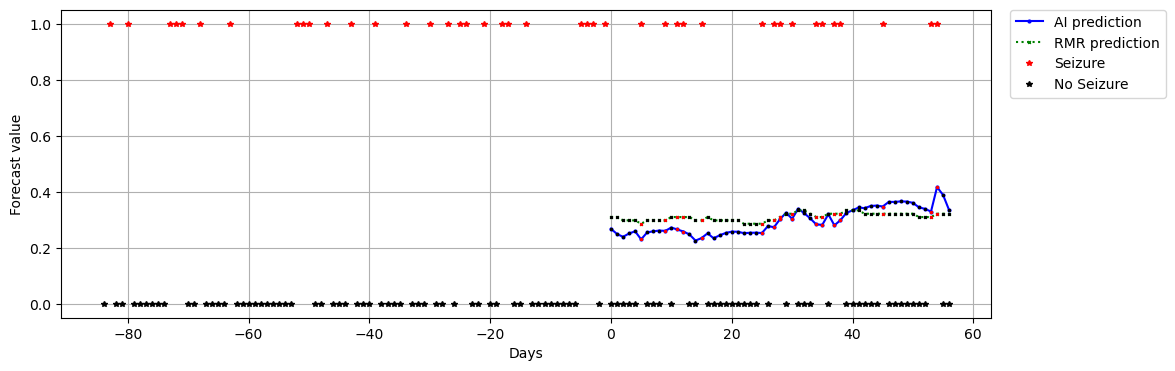

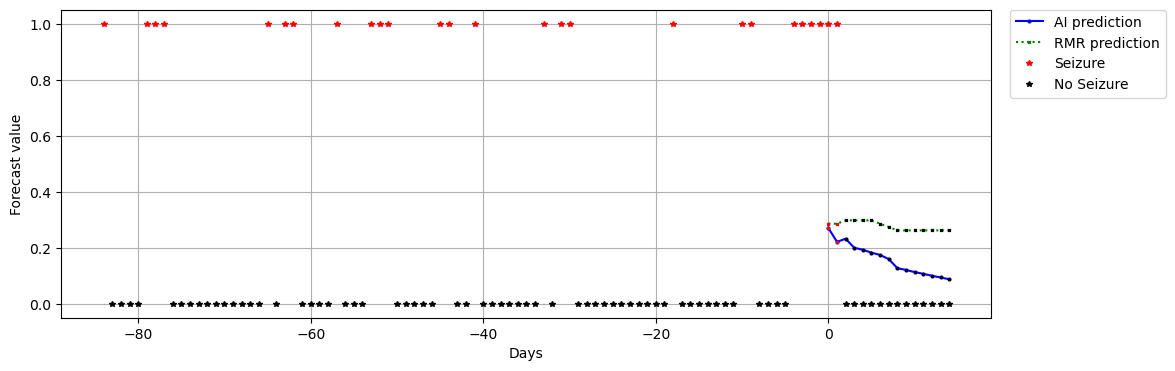

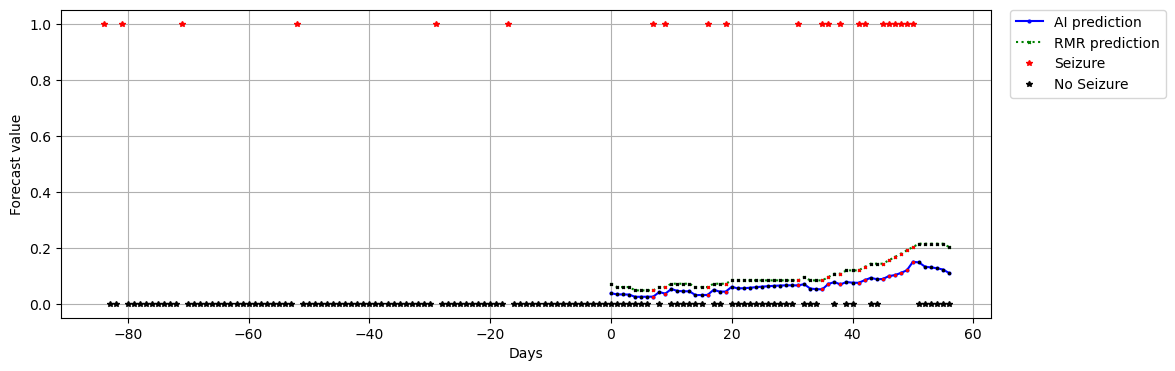

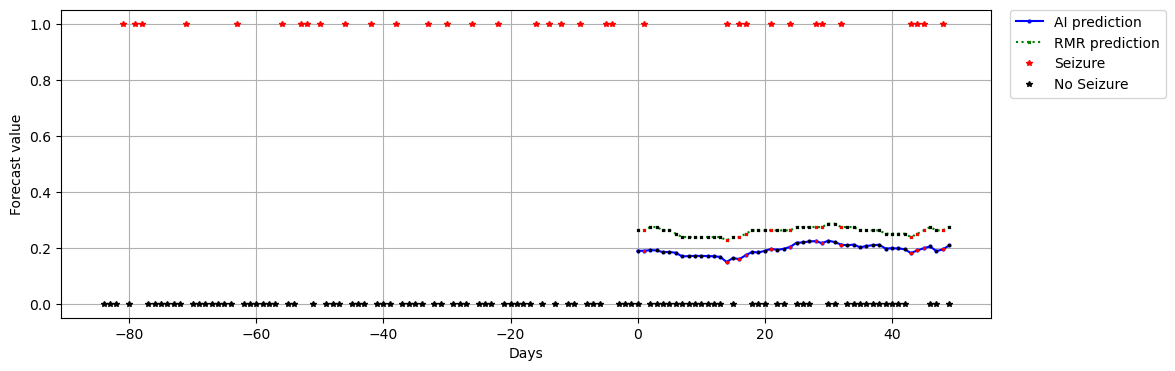

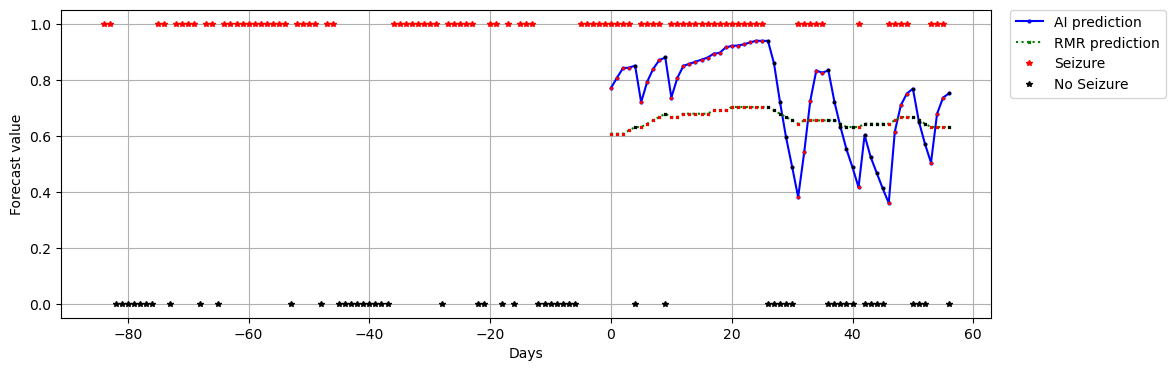

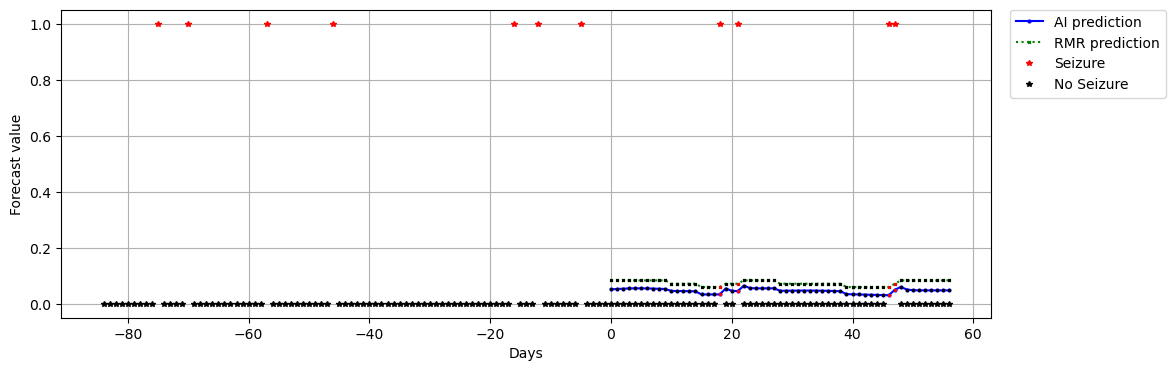

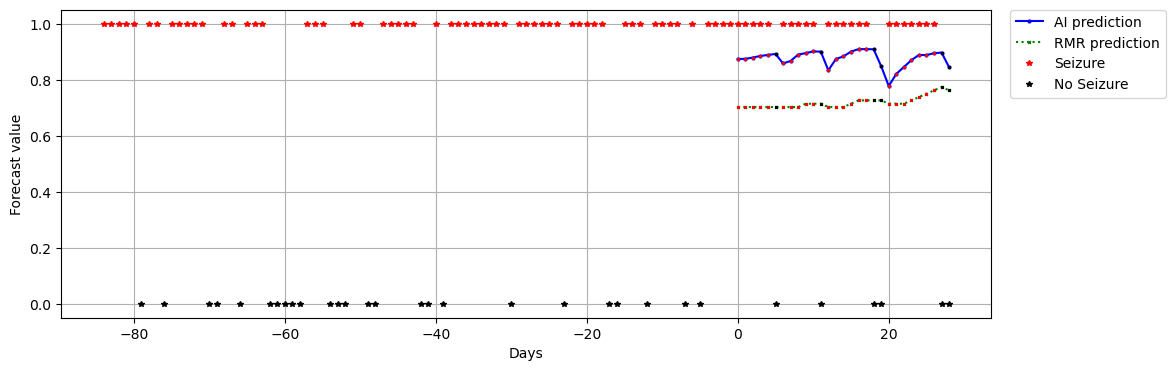

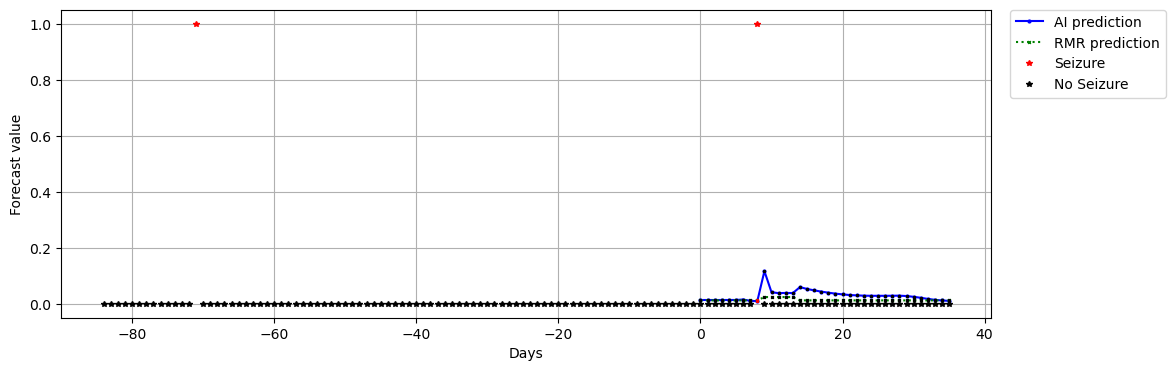

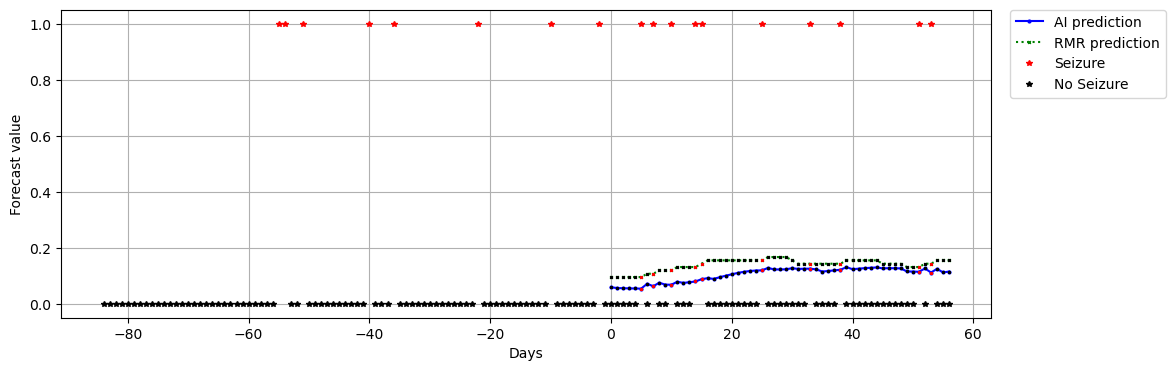

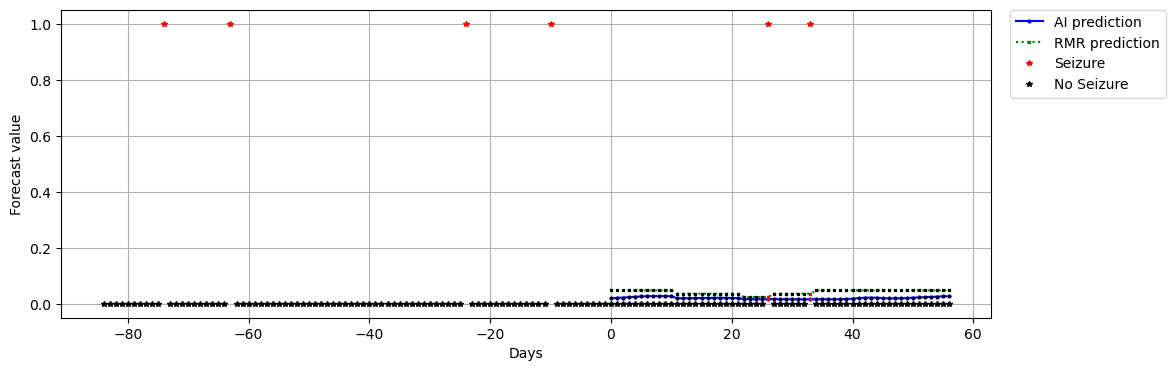

G. **Individual scores**

Unlike the AUC values in the main manuscript, which represent sensitivity versus false negative across all patients, the individual AUC scores were computed by plotting the ROC of sensitivity versus time in warning for each patient. The individual AUC time in warning scores were 0.48 +/- 0.12. Using a bootstrapped surrogates (1000 repetitions of permuted timeseries) for each patient, the AUC time in warning from the original timeseries was higher than the other permutations <5% of the time in 1 out of 25 patients.

These results can be taken into context of other published studies. One study^3^, based on intracranial RNS and sub-scalp data and GLM and RNN models found a median AUC of 0.69-0.70 with 79%-81% of patients (n=157 RNS and n=2 subscalp) showing a form of statistically significant improvement over chance. Another study^4^ using sub-scalp electrodes reported an AUC of 0.88 in one example patient with a random forest classifier. A study (n=50) using self-reported e-diary seizures^5^ found that using a combination of slow and fast cycles to forecast risk of next seizure resulted in AUC of 0.69-0.94, with all participants having statistically higher than chance results using permutation analysis (p<0.05). The forecasts in that study used a sliding 1 hour forecasting horizon, with step size of 5 minutes, making these findings challenging to compare directly to our study of 24 hour forecast horizon with 24hr step size.

The individual Brier Skill Scores (AI vs random) were correlated with the fraction of days with seizures (r=0.854, p<0.0001). This strong correlation suggests that the key advantage of the AI occurs when the seizure rate is high, specifically when the fraction of days with seizures is 30% or higher. Twelve patients had a rate >=30% during the forecasting period. All participants that stated their initial weeks of diary were accurate used their 3 months pre-enrollment as input for the moving average and AI algorithms (Appendix C). A number of participants did not record a seizure rate during this 3-month pre-enrollment rate that was as high as their prospectively recorded rates. It appears likely that the lower rates reflect underreporting.

H. **Expectations from patients about forecasts**

The participants of this study were given an initial survey prior to the 5 months of follow up. 46 eligible patients filled out this survey – though some did not move on to complete the rest of the longitudinal study. SOME of the questions reflect the perceptions of patients with respect to seizure forecasting. Below are those questions and the percentage of responses to each answer.

“If we could prove that seizure forecasts for you were accurate, would you use them?”

a. Yes - 91%
b. No	- 0%
c. NOT SURE - 7%

“How many times would you accept an incorrect forecast of LOW risk (probably not going to happen) for seizures?”

a. Zero – 0%
b. Once a year – 4%
c. Once a month – 9%
d. Once a week – 7%
e. I would not mind – 52%
f. I don’t know – 24%

“If a seizure forecast told you that your 24-hour risk for seizures was LOW (probably not going to happen) on a certain day, what would you do? (check all that are relevant)”

a. Drive. – 7%
b. Skip medications – 0%
c. Swim or take a bath unattended - 9%
d. Use power tools – 11%
e. Other ____ - 11%
f. I would not change my behavior- 80%
g. I don’t know – 9%

“How many times would you accept an incorrect forecast of HIGH risk (probably going to happen) for seizures?”

a. Zero – 2%
b. Once a year – 9%
c. Once a month – 2%
d. Once a week – 7%
e. I would not mind – 52%
f. I don’t know – 26%

“If a seizure forecast told you that your 24-hour risk for seizures was HIGH (probably going to happen) on a certain day, what would you do? (check all that are relevant)”

a. Take extra medication – 10%
b. Avoid risky behavior (driving, etc.) – 54%
c. Other ____ - 17%
d. I would not change my behavior – 28%
e. I don’t know -17%

The initial surveys (n=46) included some questions related to seizure forecasting. A sizeable fraction (52%) of patients indicated that they would not mind poor quality HIGH RISK forecasts, and a similar fraction (52%) did not mind poor quality LOW RISK forecasts, yet almost all (91%) wanted to have access to forecasts. In the setting of LOW RISK forecasts, 80% said they would not change their behavior, yet in HIGH RISK only 28% would not change – many stated that they would avoid risk-taking behavior (54%).

I. **Permuted forecast finding**

When using the permuted forecast as a reference for the Brier Skill Score (BSS), there was a noted effect that at the individual level, the BSS was often less than or equal 0, indicating an inferior comparison to the reference. However, the same comparison when conducted at the group level was found to be vastly superior to the permuted reference. To further investigate this issue, we built “groups” from size of 1 up to size 24 (there were a total of 25 non-trivial diaries to study). The groups were obtained via bootstrapping at the patient level with replacement, and 1000 repetitions were used. Shown below are the median group level BSS values and 95% confidence intervals, with permuted AI as the reference and AI as the candidate. Groups were obtained by concatenating, treating all forecasts (across patients) as if they were from a single patient. Seen here, with increasing group size, the median BSS values increase rapidly with increasing group size, reflecting the heterogeneity of seizure rates between patients rather than some newly found skill of the AI forecasts.

Picking out: 1 bss(vP) -0.0043 +/- [-0.041, 0.19]

Picking out: 2 bss(vP) 0.12 +/- [-0.020, 0.91]

Picking out: 3 bss(vP) 0.26 +/- [-0.0090, 0.84]

Picking out: 4 bss(vP) 0.38 +/- [0.0067, 0.86]

Picking out: 5 bss(vP) 0.42 +/- [0.022, 0.82]

Picking out: 6 bss(vP) 0.42 +/- [0.041, 0.79]

Picking out: 7 bss(vP) 0.46 +/- [0.046, 0.78]

Picking out: 8 bss(vP) 0.50 +/- [0.068, 0.77]

Picking out: 9 bss(vP) 0.49 +/- [0.084, 0.78]

Picking out: 10 bss(vP) 0.50 +/- [0.10, 0.75]

Picking out: 11 bss(vP) 0.50 +/- [0.14, 0.74]

Picking out: 12 bss(vP) 0.52 +/- [0.12, 0.74]

Picking out: 13 bss(vP) 0.51 +/- [0.14, 0.72]

Picking out: 14 bss(vP) 0.51 +/- [0.18, 0.74]

Picking out: 15 bss(vP) 0.52 +/- [0.20, 0.73]

Picking out: 16 bss(vP) 0.52 +/- [0.18, 0.73]

Picking out: 17 bss(vP) 0.51 +/- [0.21, 0.71]

Picking out: 18 bss(vP) 0.51 +/- [0.23, 0.72]

Picking out: 19 bss(vP) 0.51 +/- [0.23, 0.71]

Picking out: 20 bss(vP) 0.52 +/- [0.25, 0.71]

Picking out: 21 bss(vP) 0.53 +/- [0.21, 0.70]

Picking out: 22 bss(vP) 0.51 +/- [0.25, 0.70]

Picking out: 23 bss(vP) 0.52 +/- [0.27, 0.71]

Picking out: 24 bss(vP) 0.52 +/- [0.27, 0.69]
